# A coupled electro-mechanical approach for early diagnostic of carpal tunnel syndrome

**DOI:** 10.1101/2023.06.16.23291511

**Authors:** Saveliy Peshin, Julia Karakulova, Alex G. Kuchumov

## Abstract

Carpal tunnel syndrome (CTS) is a pathology affecting hand function caused by median nerve overload. Numbness in the fingers, a loss of sensory and motor function in the hand, and pain are all symptoms of carpal tunnel syndrome. The lack of numerical data about the median nerve mechanical strain inside the carpal tunnel is the main disadvantage of current clinical approaches employed in carpal syndrome diagnostics. Moreover, application of each diagnostic method alone often leads to misdiagnosis. We proposed a combined approach including hand motion capture, finite element modelling (FEM), and electromechanical simulations to evaluate median nerve compression and find a correlation with hand mobility. The hand motion capture provided the boundary conditions for FEM. After that, FEM simulations of finger flexion and hand flexion / extension were performed. Further, FEM results were put in the electrical model of nerve conduction based on the Hodgkin-Huxley model and extended cable equation. It was exhibited median nerve conduction reduced significantly throughout the flexion and extension of the hand that compared to finger flexion. During finger flexion and hand flexion and extension, the load distribution over each of nine finger flexor tendons was evaluated. The tendons of the index finger were found to have the highest Mises stress values. It was found how tendon and connective tissue contact types affected carpal tunnel pressure. The difference between the contact types was 31.7% for hand extension and 59.9% for hand flexion. The developed approach has the potential to become an alternative diagnostic method for CTS at early stages. Additionally, it can be employed as non-invasive procedure for evaluation of carpal nerve stress.

## Introduction

Carpal tunnel syndrome (CTS) is the most common tunnel neuropathy [1–3]. The loss of sensory and motor functions in the hand impairs personal and social functioning [4]. CTS patients were found to be challenged by lack of hand mobility and failure to perform basic daily activities such as holding a pen, carrying out a job, or having a healthy night’s sleep [5]. Initially, CTS only causes numbness and pain. The mild sensations can be rapidly eliminated by shaking out your hand. Rarely do people seek treatment in this situation. But as the suffering worsens day by day, sufferers feel compelled to get help. Additionally, every extra day of waiting increases the likelihood of serious nerve damage, which will make a person disabled. Meanwhile, hand pain is not always carpal tunnel syndrome [6,7]. Relevant diagnostic methods at the moment are: questionnaires, movement tests, electrodiagnostic studies, magnetic resonance imaging (MRI), computer tomography (CT) and ultrasound investigation [8]. It should be noted that these diagnostic techniques’ specificity and sensitivity are not always great, particularly at the early stages of CTS diagnosis. Additionally, utilizing just one method alone frequently results in incorrect diagnosis [9,10].

### Questionnaires

Mononeuropathy clinical guidelines include questionnaires in many countries [11]. The most informative ones are: Boston Carpal Tunnel Questionnaire (BCTQ) [12], Disability of the Arm, Shoulder, and Hand Outcome Measure (DASH) [13], Michigan Hand Outcome Questionnaire (MHQ) [14], Upper Extremity Functional Scale (UEFS) [15] and others [16–19]. The stage of the disease can be determined based on the results of the questionnaire.

It is evident that none of the outcome measures utilized in CTS patients can be regarded as the best or ideal outcome measure. Therefore, the clinician should choose the outcome measure in accordance with the main goal of their study and the target audience, the environment, and the time restrictions [20].

### Provocative tests

Sensitive and specific provocative tests are important components of the physical and diagnostic examination of human’s body. The most commonly used provocative tests for CTS are Tinel sign, Phalen’s test, and Durkan’s test [21]. On the one hand, these methods can be used equally with questionnaires to speed up the general understanding of a person’s health status [22]. On the other hand, these methods are not completely relevant for making a diagnosis, especially in the early stages of carpal tunnel syndrome.

### Injection tests

Another diagnostic test is the glucocorticosteroid injection into the carpal tunnel area. If the patient reacts positively afterwards (pain disappears), CTS is considered confirmed [23]. However, patients may decline treatment, if they experience anxiety after receiving a glucocorticosteroid injection [24].

### Electrodiagnostic testing, MRI and ultrasound studies

Electrodiagnostic studies include nerve conduction studies and electromyography. Nerve conduction studies confirm CTS by detecting abnormal median nerve conduction through the carpal tunnel when conduction is normal elsewhere [25]. The most clinically informative indicators are the M-response amplitude and median nerve conduction velocity. Electrodiagnostic studies have a sensitivity of 56% to 85% and a specificity of 94% to 99% for CTS [26]. Results may be normal in 30% of patients with mild CTS [27]. In addition, MRI is commonly used in CTS diagnosis as an objective method of determining the extent of morphological changes in the carpal tunnel [28]. However, MRI is difficult to assess the severity of the clinical course of the syndrome or the condition of the median nerve over time. Changes to the median nerve can be assessed not only by MRI, but also by ultrasound [29,30]. Ultrasound can detect the following characteristics: an increase in the cross-sectional area of the median nerve at the level of the carpal bones (distal carpal fold) > 9.8 mm^2^ [31], flattening of the nerve at the entrance to the carpal tunnel [32], echogenic features variations [33] and estimate the transverse sliding patterns of the median nerve during finger movements [34]. Electrophysiological tests do not seem to be as cost-effective as using diagnostic ultrasonography to diagnose CTS [35].

### Finite element modelling

Numerical values for carpal tunnel pressure can be determined using the finite element modelling (FEM). This method is widely used in various fields and has recently been applied in medicine [36]. The human carpal tunnel area includes tendons, ligaments, muscles, bones, nerves and others [37]. Thus, the application of the FEM was focused on studying the stress-strain state of all these tissues [38–49]. The transverse ligament is the upper arch of the carpal tunnel. It holds the tendons inside the canal and protects the median nerve from external loads. In this context, the studies by Guo [39] and Yao [44] have aimed at determining the load on the transverse ligament. Ko and Brown [38] investigated the effect of fluid within the carpal tunnel on median nerve stress using FSI calculations. Their study shows that the effect of fluid is low compared to the mechanical contact of the tendons. The subsynovial connective tissue (SSCT) is inside the carpal tunnel and encompasses the tendons and median nerve. Biomechanical approaches using the FEM were developed by Henderson et. al. [42] and Chang et. al. [45] to determine SSCT mechanical behavior. The carpal bones are the lower arch of the carpal tunnel. On the other hand, the carpal bones have mobility during hand flexion and extension. Thus, the FEM has been widely applied to model the carpal bones [41,47–49]. Perevoshchikova et. al. [41] considered a 3D-printed scaffold for scapholunate ligament reconstruction. It is also good to look at the studies where researchers have created complex models of the carpal tunnel, tendons and fingers [40,43,46,50]. Mouzakis et. al. [46] developed a model based on a slice of an MRI scan. The model includes all tissues occurring in the carpal tunnel. Wei et. al. [40] also reconstructed a 3D model based on MRI images. In their study, the carpal bones and the bones of all the fingers of the hand were considered. In this way, a model of the entire hand was created. Lv et. al. [43] also developed a complex 3D finite element model based on MRI images. Their geometric model included 29 bones, such as 14 phalanges, 5 metacarpals, 8 carpal bones and parts of the flexor pollicis longus, extensor pollicis longus, extensor pollicis brevis, extensor indicis and extensor indicis minimi; ligaments, such as the extensor retinaculum, flexor retinaculum, and annular ligaments that act as finger pulleys at the Interphalangeal (IP) joints and Metacarpophalangeal (MCP) joints ulna and radius; 9 muscles and their tendons, such as the FDS, FDP, ED, flexor pollicis brevis. In order to successfully implement finite element calculations, these studies use Young’s modulus and Poisson’s coefficient for the tissue simulation. However, the mechanical properties of the soft tissues within the carpal tunnel have also been studied in several studies.

An important aspect of FEM is the mathematical model that describes the mechanical behavior of the tissue. At the same time, any model chosen requires a set of mechanical parameters for each tissue. For example, describing the behavior of tissue by a linear elastic equation requires two parameters: Young’s modulus and Poisson’s coefficient. This approach is convenient for calculations since it requires less computational resources. However, other approaches are used to obtain more accurate and valid results. Osamura et al. [51] defined the overall mechanical response with linear-elastic material properties; however the data curves appear to be non-linear with a strain-dependent response [52]. Main et el. [53,54] identified the tendon and median nerve properties as isotropic, non-linear, hyperelastic and described their mechanical behavior with Ogden model [55] and determined the parameters α and μ. Our previous work [50] also included modelling of the entire hand, including tendons, carpal tunnel, carpal bones and phalanges. At the same time, soft tissue materials have been defined by the Ogden model with parameters from Main et al. study. However, the carpal tunnel has been simplified and fixed, which corresponds to finger flexion, but does not accurately correspond to hand flexion. In this study, this lack will be taken into account in the FEM.

### Nerve conduction modelling

The median nerve contains sensory, motor and autonomic fibers. Sensory nerve fibers are known to lose their conductivity faster than motor fibers if the nerve is under external mechanical stress [56–59]. The classical mathematical model describing the conduction of an electrical signal along the axon of a neuron was developed by Hodgkin and Huxley [60]. The model is based on differential equations system that describe the membrane exchange of *Na* and *K* ions. Further, FitzHugh [61] introduced a simplified Hodgkin-Huxley model (H-H), which consists of two equations versus Hodgkin-Huxley’s order of twenty. Later, these models have been improved and refined by other researchers, but the basic approach remains the same. However, these models describe the action potential at a single point. In order to simulate median nerve conduction, it is necessary to describe its conduction along a defined distance. The traditional cable equation is used [62] for the action potential propagation along the neuron. The extended cable equation is used to take into account the shape effect of the peripheral nerve. Changing the cross-sectional area of the nerve reduces conduction. Coupling the conduction model and the cable equation is used to describe nerve fiber conduction [63–68].

### Aim of study

The aim of this study is to develop a combined electro-mechanical approach for early diagnosis of carpal tunnel syndrome based on modern biomechanical and mathematical modelling methods as well as applying the developed approach and obtaining new numerical results in the stages of the approach.

## Materials and Methods

This paper presents a combined electro-mechanical approach for early CTS diagnosis, including hand motion capture, FEM of the patient-specific hand based on MRI, CT, and mathematical modelling of nerve conduction (Fig 1). An important criterion of this approach is the confirmation of the disease at an early stage. This is necessary to reduce rehabilitative time and avoid traumatic surgery.

**Fig 1.**
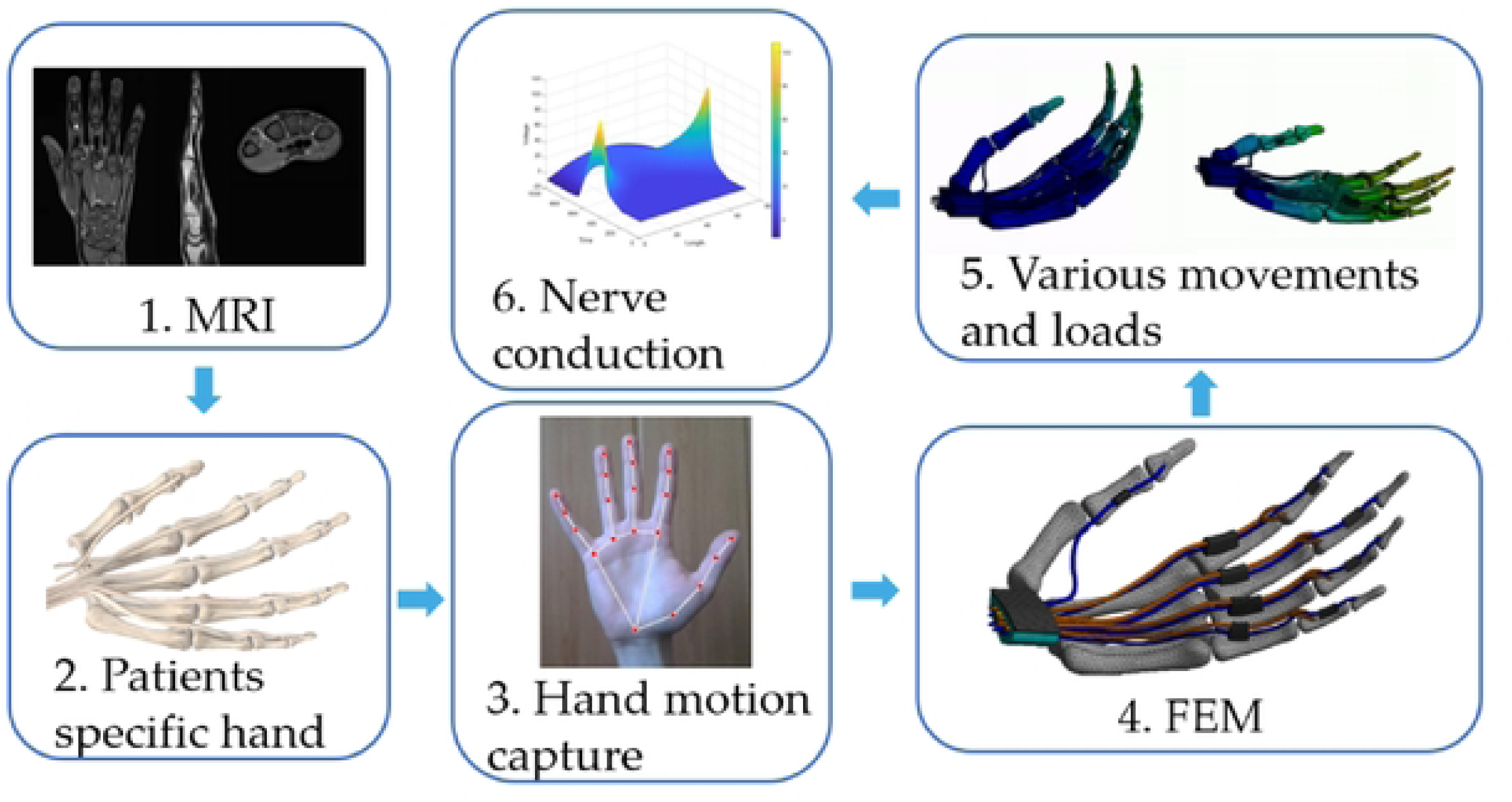
A six-stage approach for the early diagnosis of carpal tunnel syndrome.

### Data collection

The first stage is performing an MRI and CT scan of the volonteer’s hand. The research protocol used in this study was approved by the Ethical Committee Board of Perm State Medical University (Protocol No. 17 on 25 November 2022). The volunteer had no hand disease or associated neurological disorders. Participant signed written informed consent. The participant was recruited for research purposes in the period between 5 December 2022 and 23 December 2022. Authors had no access to information that could identify individual participant during or after data collection.

DICOM files are using to create patient-specific 3D geometry of the carpal tunnel, tendons, ligaments, median nerve, and phalanges. CT scans are the best way to visualize a human’s bones. In such a case, the carpal and metacarpal bones, as well as the distal, intermediate, and proximal phalanges, were built up from CT scans. At the same time, soft tissues such as the median nerve, tendons, ligaments and connective tissue may be visible on MRI scans.

### 3d Model extraction

However, the resulting 3D models require post-processing. The post-processing of the tissues was performed using the Meshmixer software. The three-dimensional tendon geometry was built based on a set of cross-sectional planes of the obtained DICOM file geometry. The resulting personal geometry of the left hand and carpal tunnel after processing is shown in (Fig 2).

**Fig 2.**
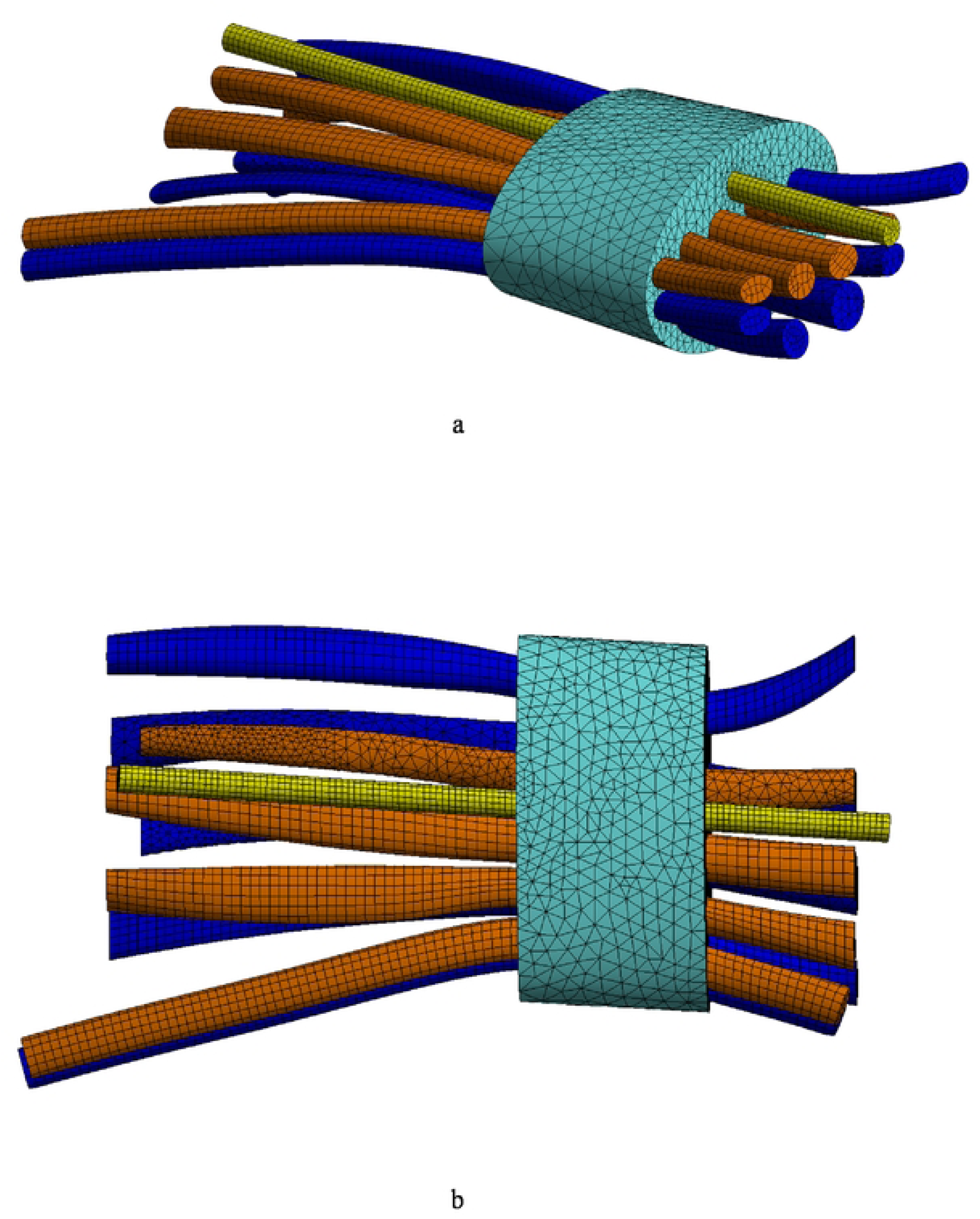
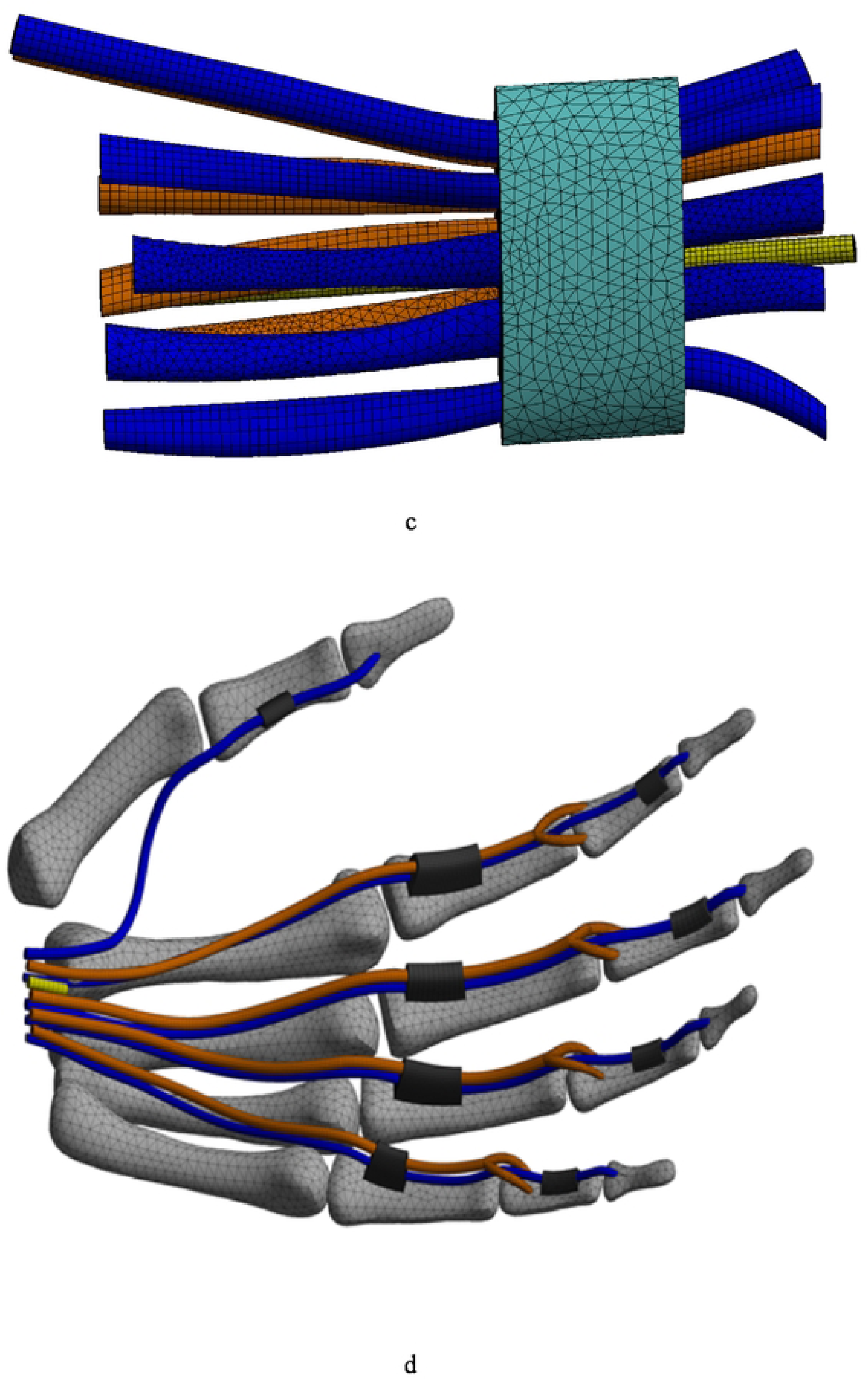
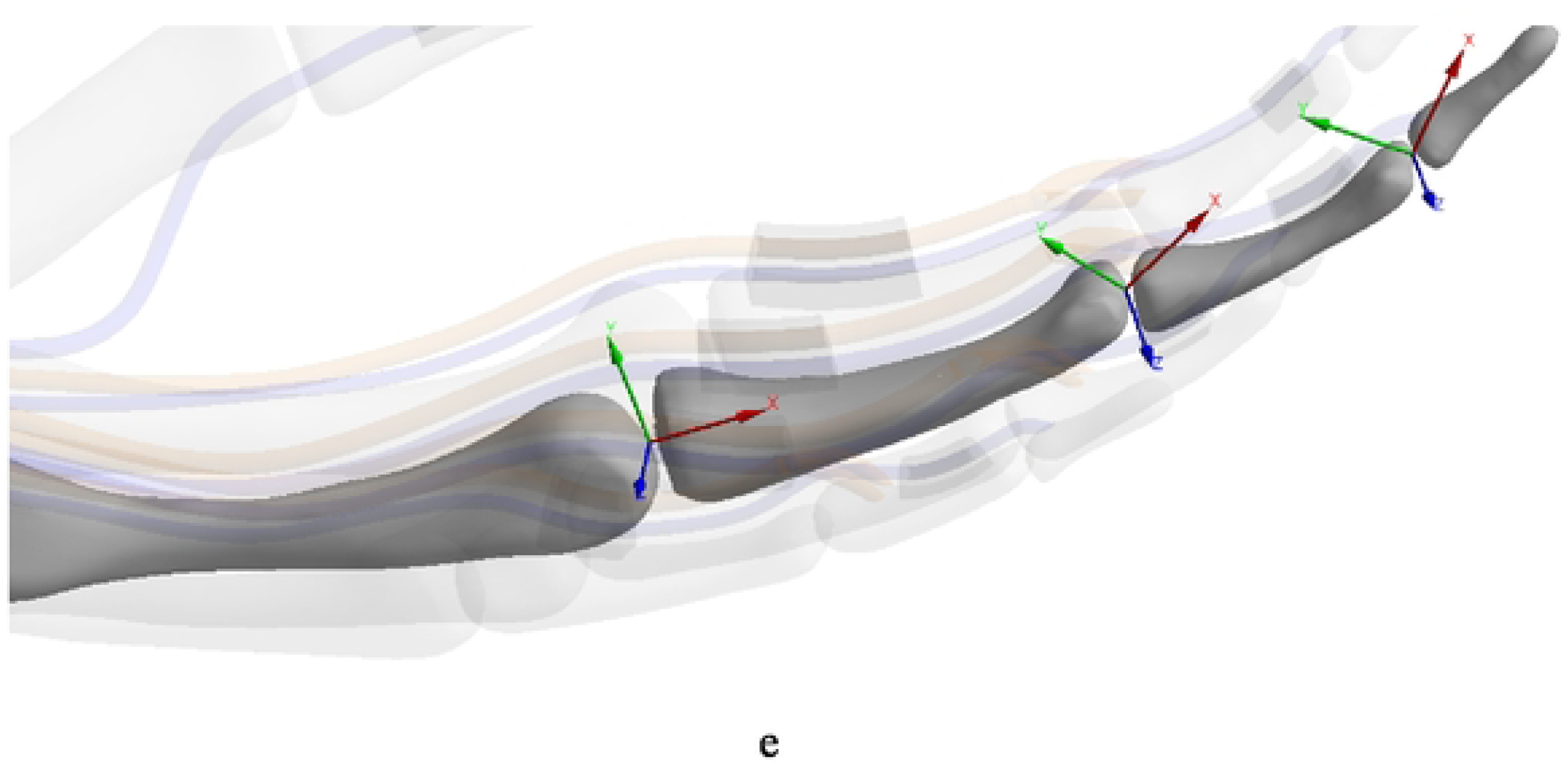
Patient-specific finite element model of carpal tunnel (a) – general view, (b) – top view, (c) – bot view and. (d) – Finite element model of a hand (carpal tunnel is hidden), (e) – The connection of the phalanges with the joint and the coordinate system of the joint.

### Hand motion capture

The human hand is a unique manipulator that can handle many mechanical tasks. Different people may perform certain mechanical tasks differently in their daily lives. For example: how to hold a fork or a mug, how to type on a computer keyboard, or how to hold a phone. Depending on age, anatomy, and gender, people can flex their fingers and move their hands differently. To take individual hand and finger movements into account, software was developed to capture hand movement in real time and determine the coordinates of characteristic hand points (Fig 3) [69]. Visual Studio Code and Python were used for software development. Learned free artificial intelligence is used in the software to determine points on the hand using video from a camera. The software applies the points to the hand, determines the coordinates of the points, and moves the position of the points during hand movement in real time. A line simulating the phalanx is drawn on the two coordinates. The angle between the two lines describing the angle between the two phalanges of one finger is then determined. Angle changes in the distal interphalangeal joint, proximal interphalangeal joint and metacarpal phalangeal joint of all fingers were obtained using the developed software according to the formulas:

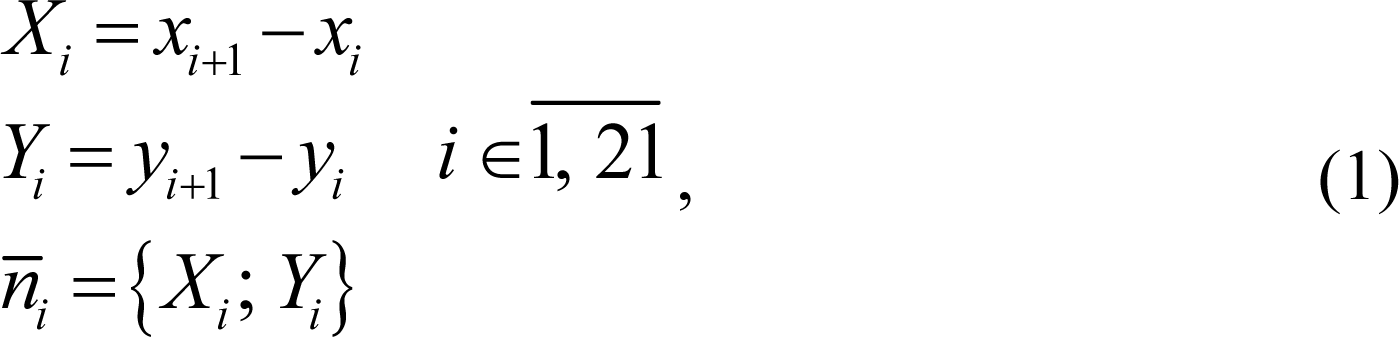

**Fig 3.**
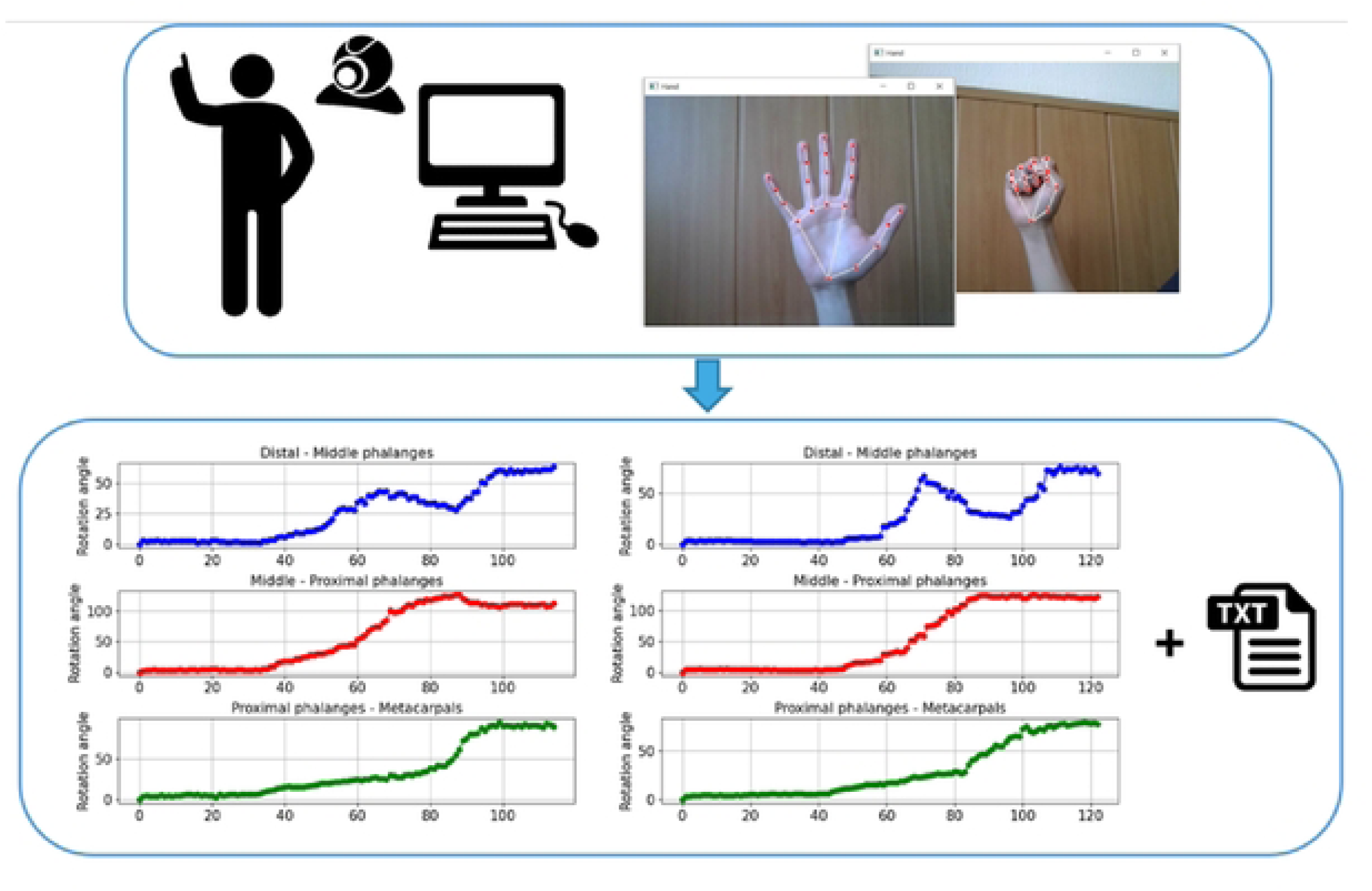
Motion capture process

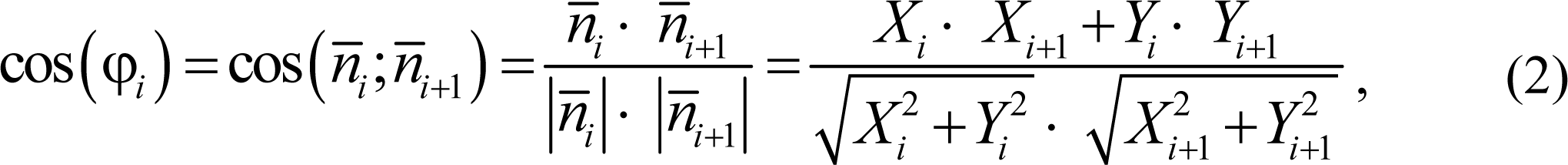

where *x*, *y* – characteristic point coordinates, *X*, *Y* – components of the normal vector *n*, *i* – characteristic point number, φ - angle between the phalanges.

### Finite element model

The fourth stage is FEM based on a patient-specific geometrical model. The FEM was used to simulate various hand and finger movements. The three-dimensional geometry was imported into the finite element software ANSYS to generate the finite element model with interaction material parameters, boundary conditions and loading conditions defined. Two types of geometry were built based on MRI and CT to calculate carpal tunnel stress during finger flexion and hand flexion and extension. The first geometric model included SSCT, median nerve, nine finger flexor tendons, transverse ligament, all phalanges, metacarpal bones, and simplified carpal bones. The finger flexion geometry consisted of 40 solid bodies that were in contact and joined together. The whole first geometry is divided into 422,779 nodes and 122,486 elements by hexahedral and tetrahedral elements. The second geometric model included SSCT, median nerve and nine tendons. The hand flexion and extension geometry consisted of 11 solid bodies. The whole second geometry is divided into 126,307 nodes and 56,723 elements by hexahedral and tetrahedral elements. Hexahedral elements are used to build the tendon mesh and median nerve in both geometries.

### Joints and contact

One type of joint (Revolute) and two types of contact (Bonded and No Separation) were used in FEM. Revolute joint limit the free movement of bodies to one degree of freedom. The only motion that is possible between two jointed bodies is rotation around the Z-axis (Fig 2e). Revolute joints are used to simulate finger flexion in the joint between the distal phalanx and the middle phalanx (distal interphalangeal joint), the joint between the middle phalanx and the proximal phalanx (proximal interphalangeal joint) and the joint between the proximal phalanx and the metacarpal bones (metacarpal phalangeal joint). Bonded contact type is used to model bodies which strain, stress and displacement in the contact zone are the same. In other words, the bodies are rigidly connected to each other and cannot move relative to each other in the contact zone. No separation contact allows the bodies in the contact zone to move along in the tangential direction without friction, and prohibits movement in the normal direction. Thus, the bodies cannot move away from each other but can slide. The types of solid contact are shown in (Table 1).

**Table 1.** Solid bodies contact types.

| **Solid body pair** | **Interaction / Contact type** |
| --- | --- |
| Distal phalange to Middle phalange | Revolute |
| Middle phalange to Proximal phalange | Revolute |
| Proximal phalange to Metacarpal | Revolute |
| Deep flexor tendon to Distal phalange | Bonded |
| Superficial flexor tendon to Middle phalange | Bonded |
| Superficial and Deep flexor tendon to Annular ligament | No separation |
| Superficial and Deep flexor tendon to SSCT | No separation / Bonded |
| Median nerve to SSCT | No separation / Bonded |
| Annular ligament to phalanges | Bonded |
| Transverse carpal ligament to SSCT | Bonded |

The median nerve was connected to the connective tissue by no separation contact type for finger flexion model. The contact between the tendon and connective tissue was no separation as well. It is well established that tendons have high mobility relative to connective tissue and that the subsynovial fluid ensures minimal friction of the tendons against the connective tissue. No Separation type of contact ensures frictionless sliding of the two bodies and applies a restriction on rupture of the contact. However, during hand flexion and extension the tendons cannot always slide along the carpal tunnel without friction. The pressure between the connective tissue and the median nerve and tendon increases with flexion and extension of the hand. Thus, it is difficult to determine the friction coefficient between these tissues in vitro. To take this effect into account, finite-element calculations were performed with two types of tendons, connective tissue, and median nerve contact. Thus, the actual carpal tunnel strain values will be between the two boundary results with no separation contact type and with bonded contact type.

### Mechanical properties of the tissues

The Ogden model [55] was used to describe the mechanical properties of the soft tissues. The model was chosen after a series of studies by the researchers [52–54] that experimentally determined the mechanical properties of all tissues within the carpal tunnel. The Ogden model was also used in these studies. The choice of this approach makes the calculations considerably more complicated, however, it allows to obtain results that are similar to the experimental data. thus, the soft tissues are modeled as isotropic linearly hyperelastic and the coefficients are shown in (Table 2). Bone deformation in flexion and extension of the hand is much less than soft tissue deformation. Thus, phalanges, carpal bones and metacarpal bones are modeled isotropic linearly elastic. Young’s modulus and Poisson’s coefficient of bone tissue are also shown in (Table 2) [70].

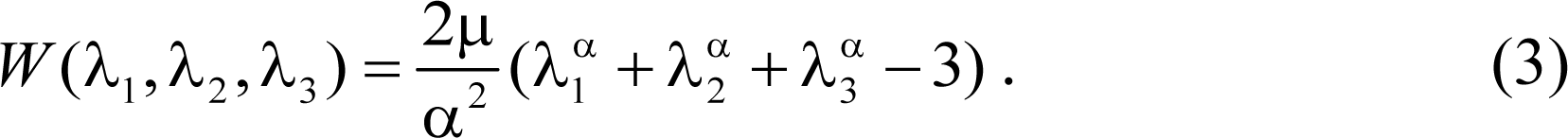

**Table 2.**
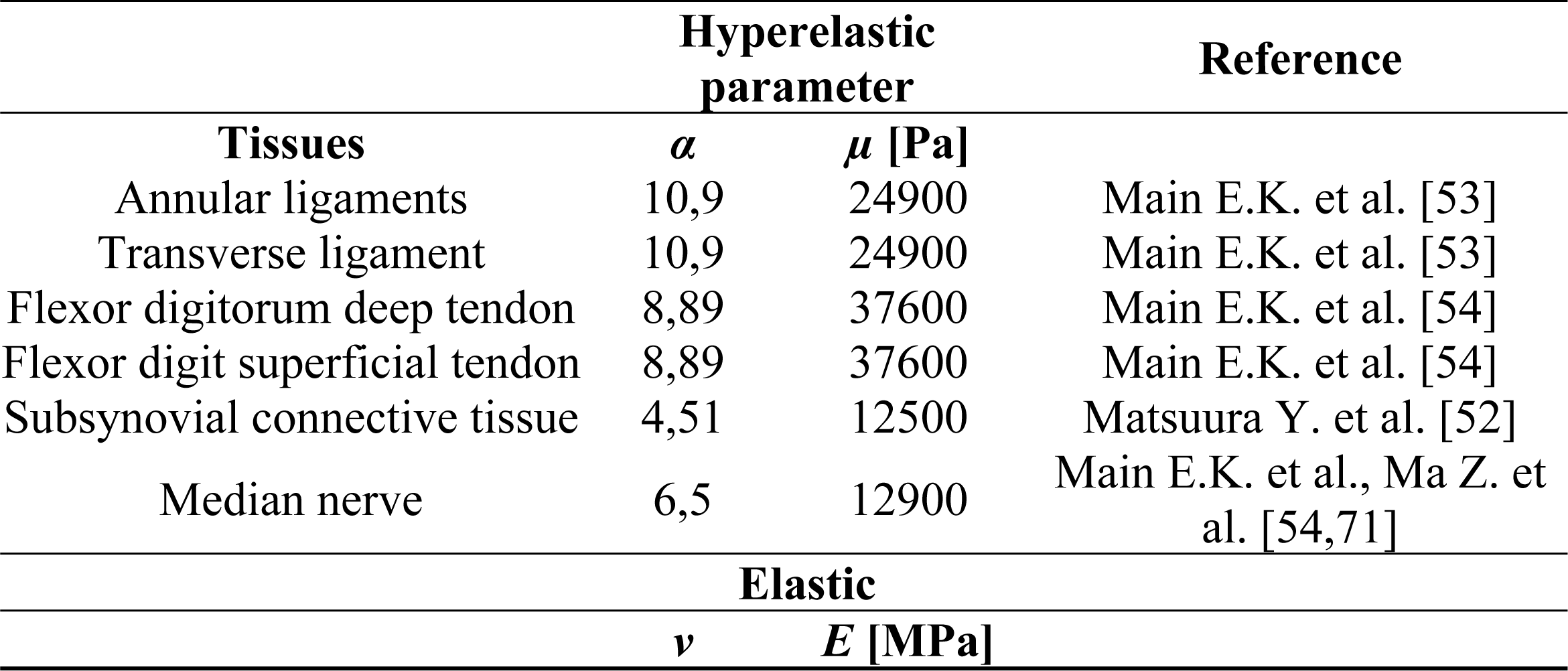
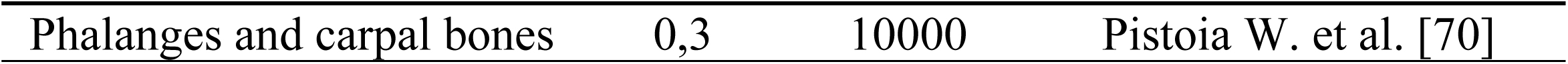
Mechanical properties of the tissues used in model.

### Boundary conditions

The different hand movements are dealt with in the fifth stage of the approach. Human finger flexion is achieved by the finger flexor muscle actions and the effort transferred via tendons to the median nerve that passes through the carpal tunnel. The load from the tendons to the median nerve is transferred by the connective tissue inside the carpal tunnel, which surrounds both the tendons and the median nerve. The movement of the tendons defines the stresses in the carpal tunnel. In these conditions, it is important to consider the actual tendon movement caused by finger flexion. For this purpose, a three-dimensional model of the finger phalanges was built (Fig 2d) and the contact between the tendons and the phalanges was defined as bonded. The carpal bones were fixed laterally, the proximal tendon surface was displaced to simulate muscle contraction, the metacarpal bones were fixed and the proximal and distal areas of the median nerve were fixed as well. The rotation angle for each pair of phalanges was set according to the results obtained during the hand motion capture stage. The angles between each pair of phalanges were determined using the software developed so far.

The human hand movement at the wrist joint is similar to the movement of a spherical cylinder [72]. This study deals with flexion and extension of the wrist at the wrist joint as well. The wrist rotation angle during these movements is maximal and therefore the median nerve compression can be large. For this, the geometry of the patient’s nine tendons, the median nerve and the connective tissue between them were reconstructed (Fig 2 a-c). To simulate hand flexion and extension in the wrist, a coordinate system attached to the geometric center of the carpal tunnel was chosen. A rotation of 30 degrees around the Z-axis of the distal tendon surfaces and the medial nerve was applied. The direction of the Z axis was reversed for hand extension simulation.

### Nerve conduction

The final stage of the approach is to determine the conduction of the deformed median nerve. When CTS occurs, the median nerve is compressed by the surrounding tissue and deformed. Nerve compression is the main cause of the CTS symptoms. Sensory and motor fibers are damaged as a result of compression. As a result, nerve fiber conduction is decreased or lost. The studies describe approaches to mathematical modelling of nerve conduction. The Hodgkin-Huxley (H–H) model [60], which describes the potential difference in the membrane potential of the giant squid axon, is considered a fundamental approach in this area. The model describes the Na and K ions behaviors, which produce the resting potential and the action potential. The model consists of a set of differential equations and is solved by the finite difference method. The distribution of the electrical signal in the cylinder is defined by the cable equation [62]. The extended cable equation gives consideration to the varying geometry of the cylinder.

### Hodgkin and Huxley model

The H–H model was developed in 1952 as a result of Hodgkin’s and Huxley’s extensive studies of the giant axon of the squid. It describes how the action potential is initiated and how it propagates in a neuron [60]. In the H-H model, the nerve cell membrane is regarded as a flat capacitor. The potential difference on the coils of the capacitor *U* is related to the stored charge *Q* and the total electrical capacitance of the membrane *C* by the relation:

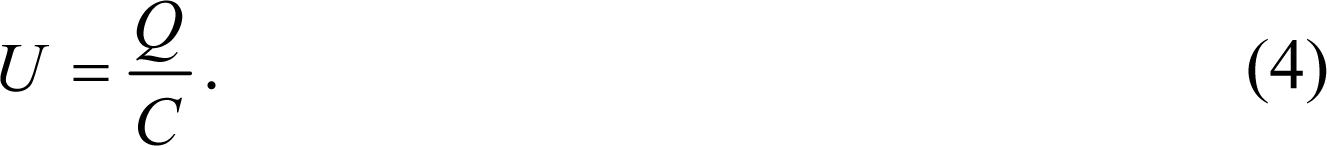

By differentiating this equation by time and denoting *dQ/dt* as the current flowing through the membrane, we obtain

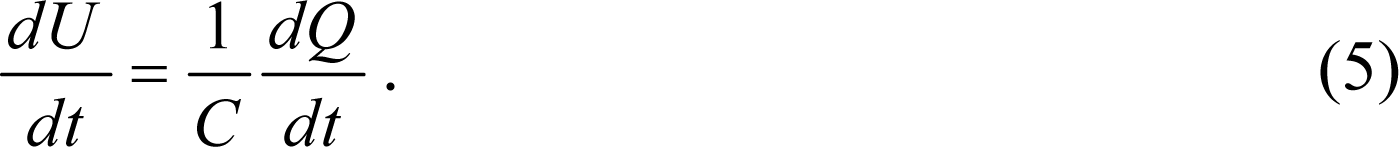

By replacing *U* by the membrane potential *E_m_* and replacing the total capacitance by the capacitance divided by the unit area and given that the main contributors to the generation of the nerve impulse are the potassium ion currents *i_K_* and the sodium ion currents *i_Na_*, we can simplify the equation:

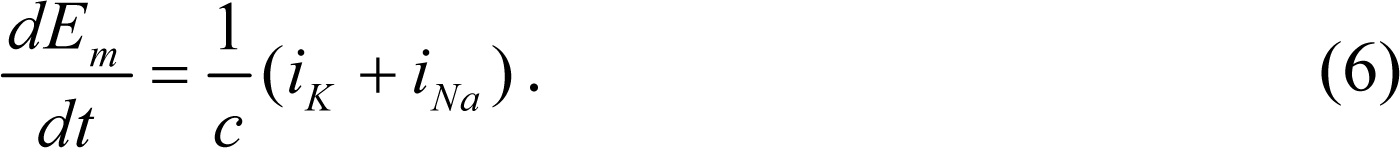

To solve the equation, it is necessary to find the dependence of specific currents through sodium and potassium ion channels on membrane potential. The relationship between electric current and potential can be described by the following equations:

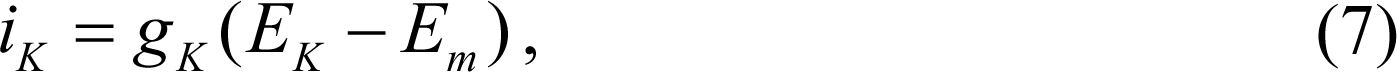

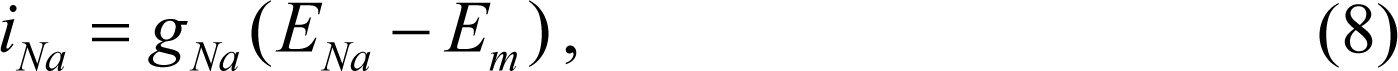

where, *g_K_* and *g_Na_* are not constants but functions of the membrane potential, which is due to the potential-dependent properties of the ion channels. In Hodgkin and Huxley, the dependence functions of *g_K_* and *g_Na_* on membrane potential have been determined on the basis of the analysis of the form of the experimental currents-time dependences under different constant membrane potential values. Hodgkin and Huxley showed that *g_K_* and *g_Na_* can be described by the equations:

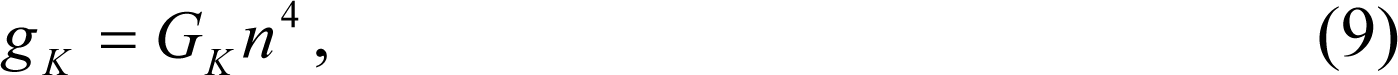

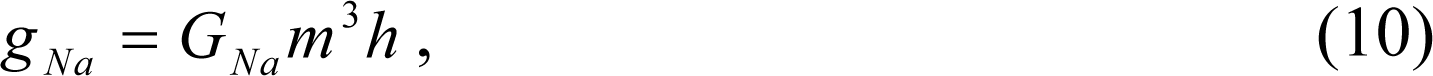

where *G_K_* and *G_Na_* are maximum conductivities for potassium and sodium channels, *n* and *m* - activation gate variables for potassium and sodium channels; *h* - inactivation gate variable for sodium channels.

The dynamics of gateway variables can be described by equations:

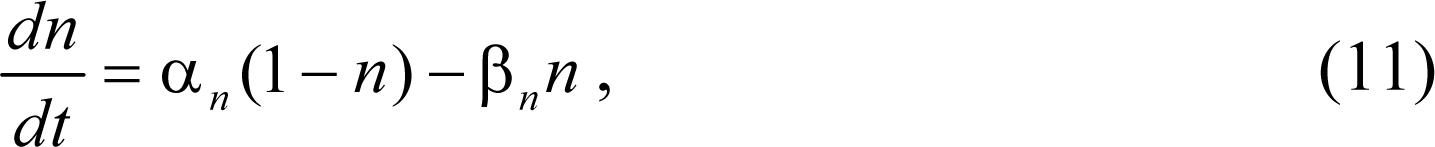

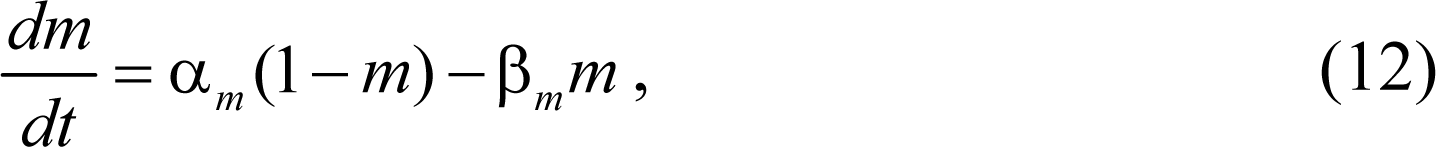

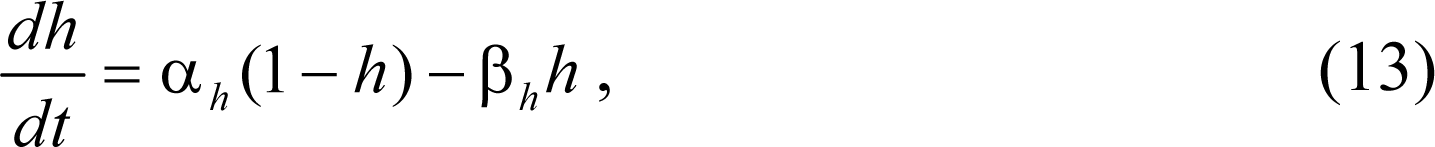

where α and β are the rate parameters for the transition of the gate particle to the «open» and «close» state, respectively. These constants also depend on the membrane potential and are described by equations:

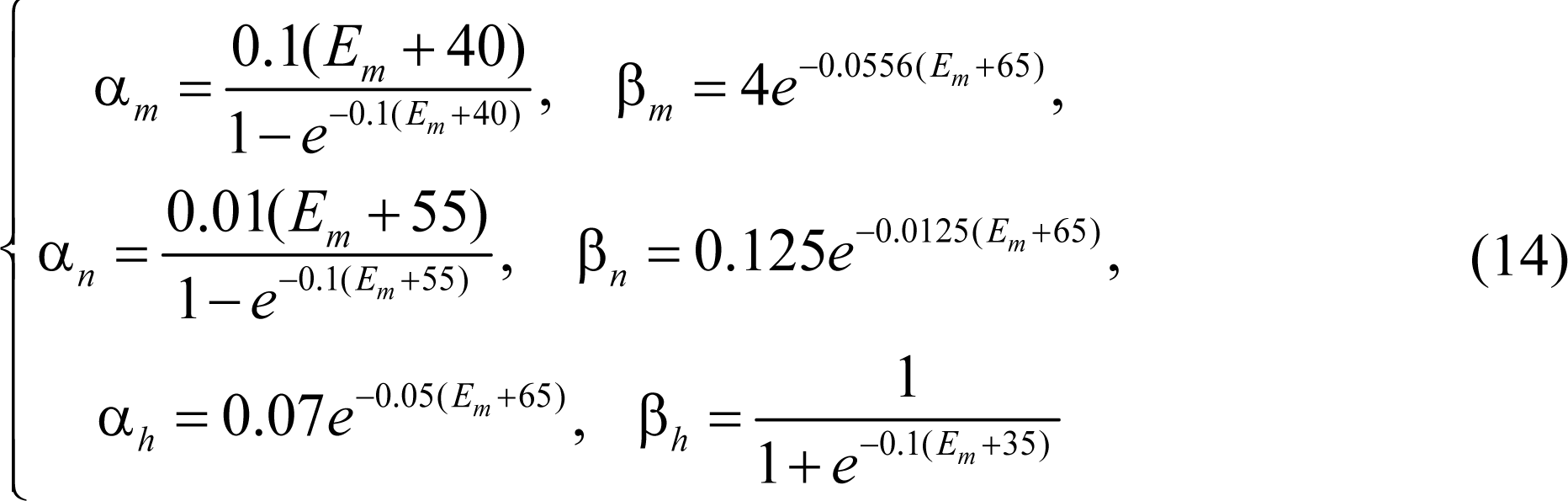

The system of equations (5)-(11) is a H-H model and is solved using a self-developed code in Matlab using the finite difference method.

### Cable equation

The cable theory is given as a second-order partial differential equation:

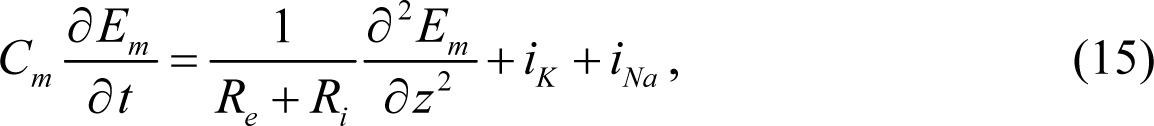

where *R_e_* and *R_i_* are extracellular and intracellular axial resistivity, respectively. The parameters used in the nerve conduction model are listed in (Table 3).

**Table 3.** Parameters used in nerve conduction model.

| Symbol | Model parameters | Value |
| --- | --- | --- |
| $E_{Na}$ | Sodium potential | 52.4 mV |
| $E_K$ | Potassium potential | -72.1 mV |
| $g_{Na}$ | Sodium conductance | 120.0m mho/cm<sup>2</sup> |
| $g_K$ | Potassium conductance | 36m mho/cm<sup>2</sup> |
| $C_m$ | Membrane capacitance | 1.0 μF/cm<sup>2</sup> |
| $R_i$ | Resistivity of intracellular space | 35 Ω cm |
| $R_e$ | Resistivity of extracellular space | 20 $\Omega$ cm |

### Extended Cable Equation

The effect of changes in nerve geometry on action potential propagation is accounted for by the extended cable equation.

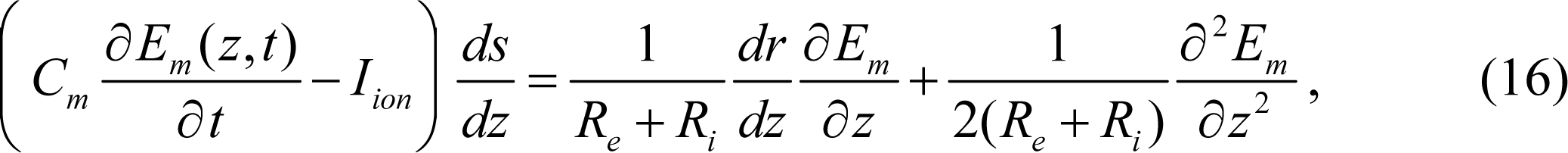

where, *r*, *z* and s are coordinate variables describing the nerve shape and *r (z) = A-B_z_-C ^2^ -* the function describing the change in the shape of the neuron. Parameters *A, B*, and *C* are parameters taken from the results of finite-element modelling. The set of equations is solved by the finite difference method in the Matlab-based code. Conduction modelling was performed on three millimeters of the median nerve in the area of highest stress.

## Results

An MRI and CT of the left hand and carpal tunnel was performed. The results obtained in DICOM files were used to construct a patient-specific three-dimensional geometry of the investigated tissues (Fig 2). Finger flexion and extension were captured and processed using the software developed. Patient-specific movement of fingers and hand is necessary to set the boundary conditions objectively in FEM. The motion capture process and its results are shown in (Fig 3). The resulting time dependencies of the rotation angles between the phalanges were used in FEM to assess the effect of finger flexion on median nerve compression. Six series of hand motion captures are shown in (Fig 4). The graphs show changes in the angle of the three phalanges over time during index finger flexion. Blue shows angle change in distal interphalangeal joint, red shows angle change in proximal interphalangeal joint, green shows angle change in metacarpal phalangeal joint. The maximum distal - middle phalanges angle was 61°, maximum middle – proximal phalanges angle was 116°, maximum proximal phalanges – metacarpals angle was 101°.

**Fig 4.**
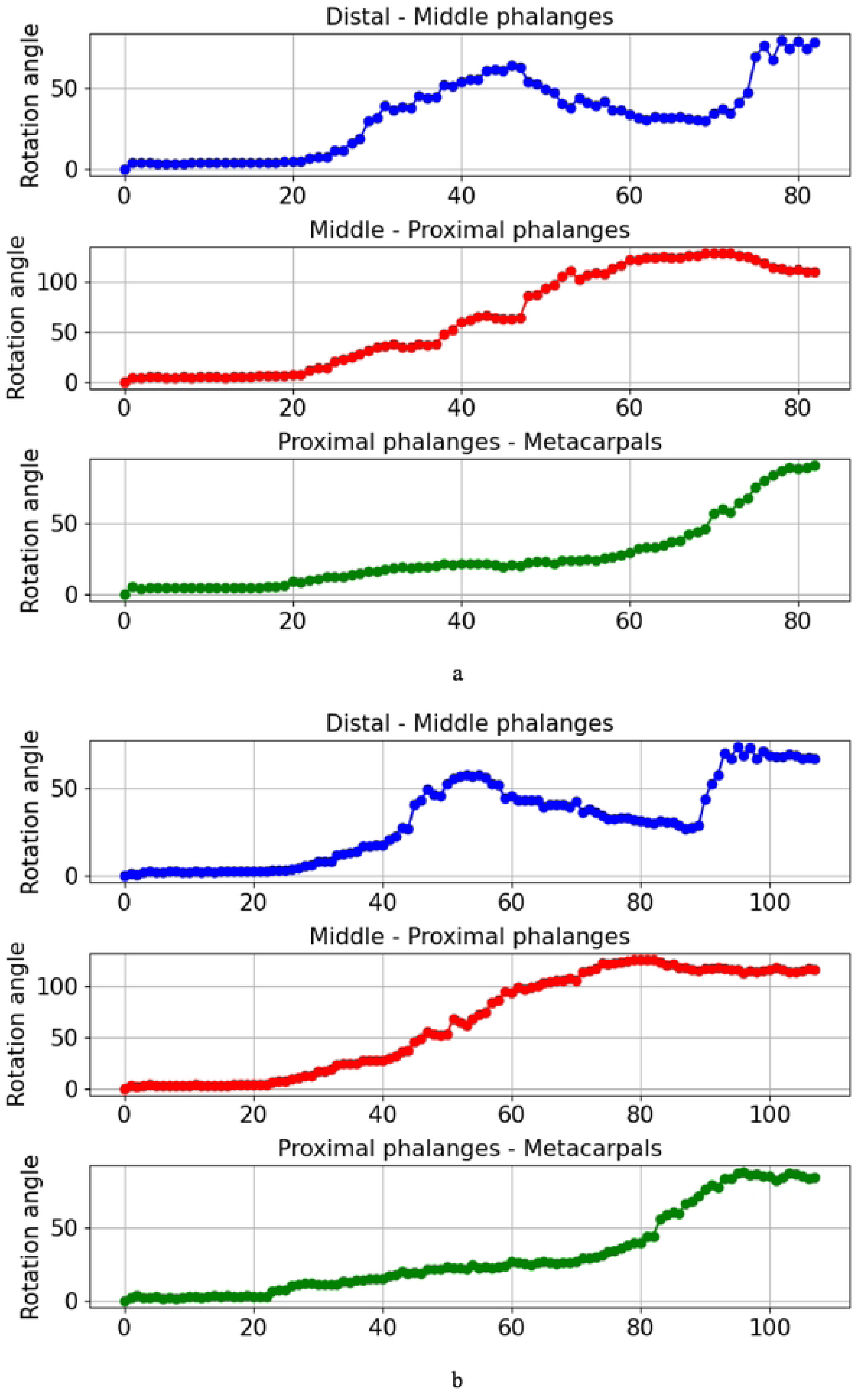
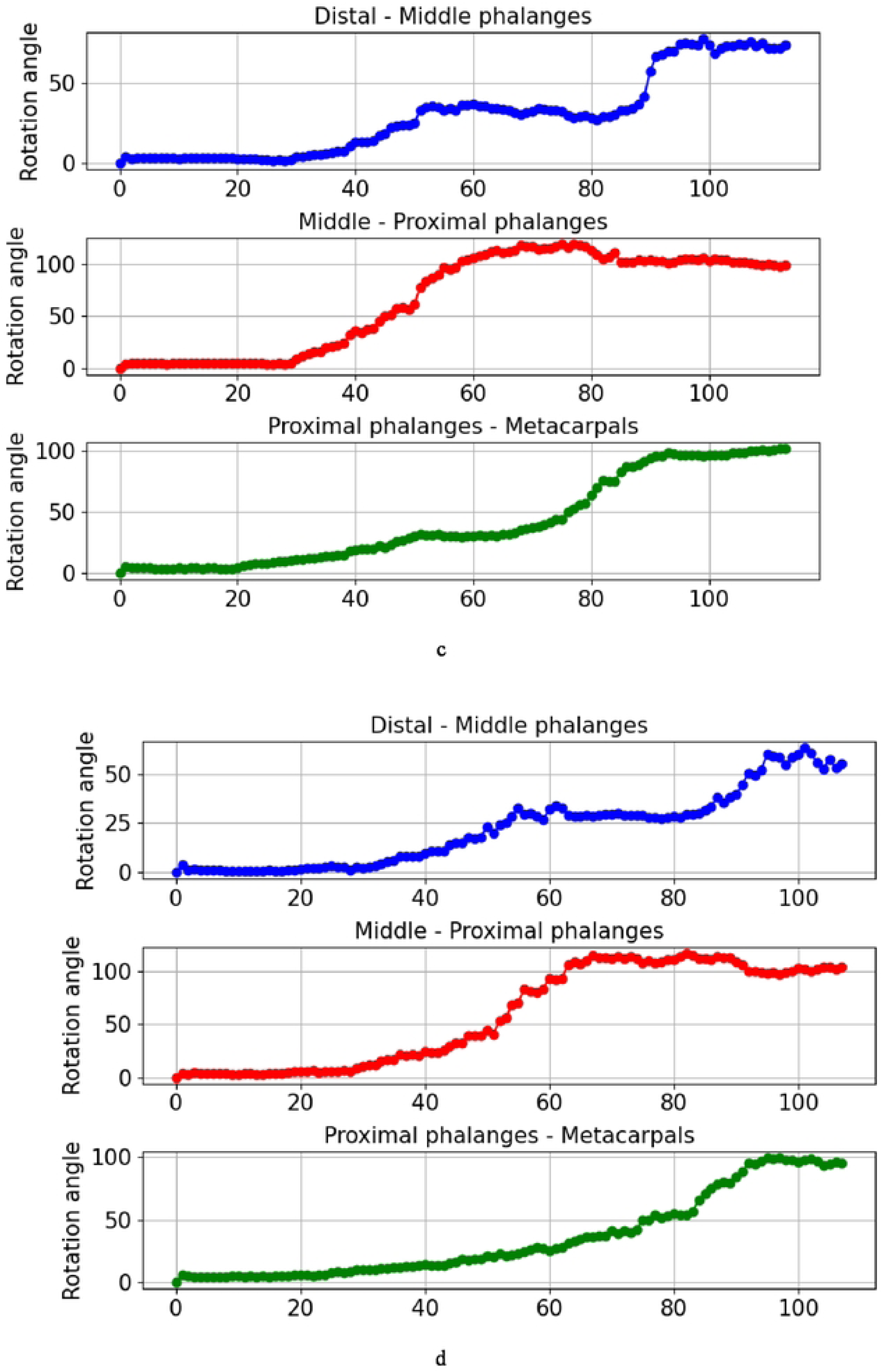
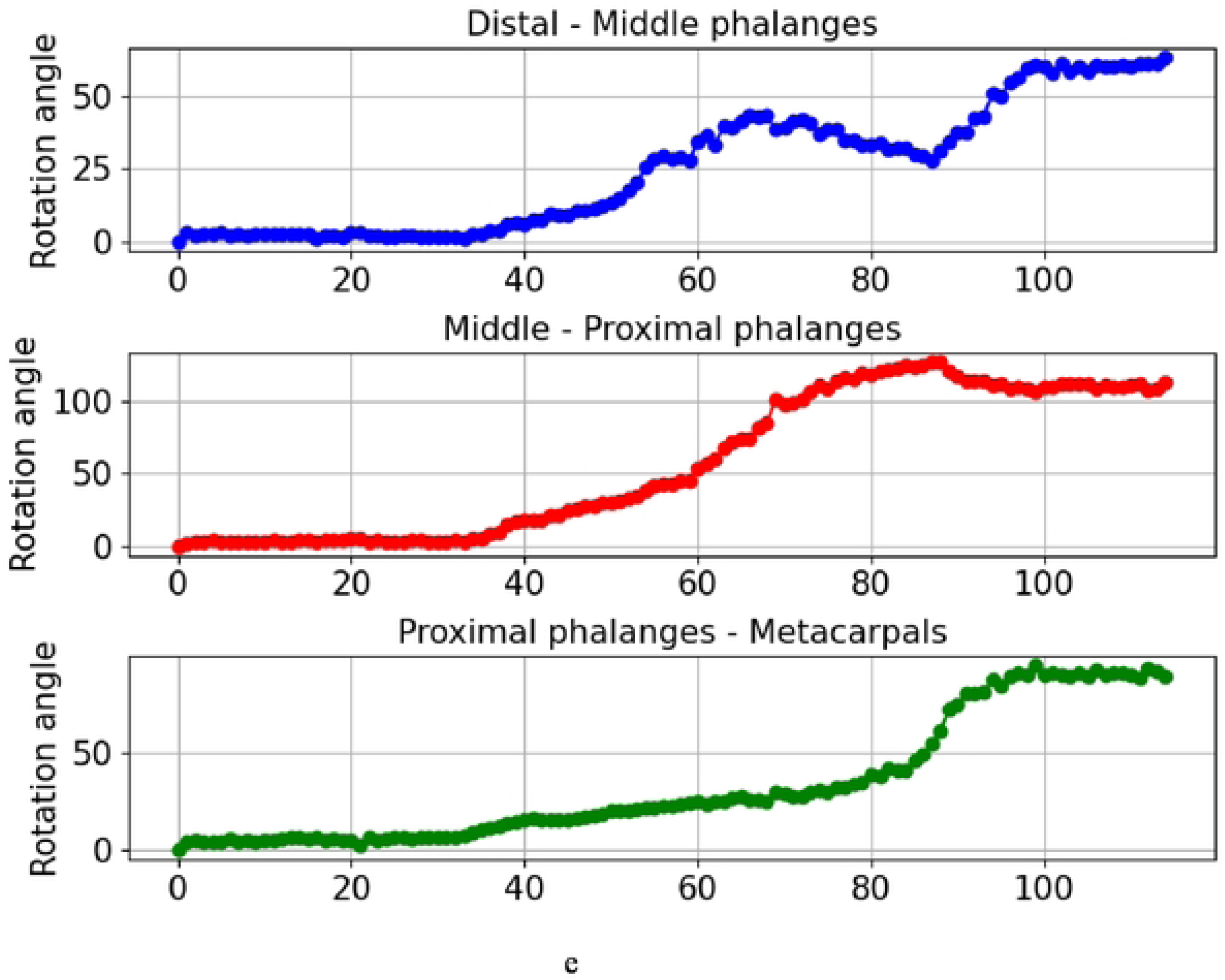
Change of angles in the three joints of the index finger over time obtained by motion capture: a-e series number from one to six.

### Fingers flexion

Stress, displacement and strain values were obtained in all modelled tissues during finger flexion. The results of the total deformations at various modelling times (*t*=3s, *t*= 15s, *t*=25s, *t*=35s) during finger flexion are shown in (Fig. 5). The maximum stresses do not always clearly describe the stress state of the soft tissues, especially close to the fixed areas. In this case, Von Mises stress along the midline of the tendon and median nerve were analyzed. The midline was constructed based on a set of midpoints of the tendon cross sections. However, the maximum von Mises stress is shown in (Table 4). Tendons were divided into two groups: deep and superficial. The thumb tendon is shown in both groups. Stress distribution along the tendon’s midline as a function of distance is shown in (Fig 6). The distal plane of the tendon is taken as the start of the distance report.

**Fig 5.**
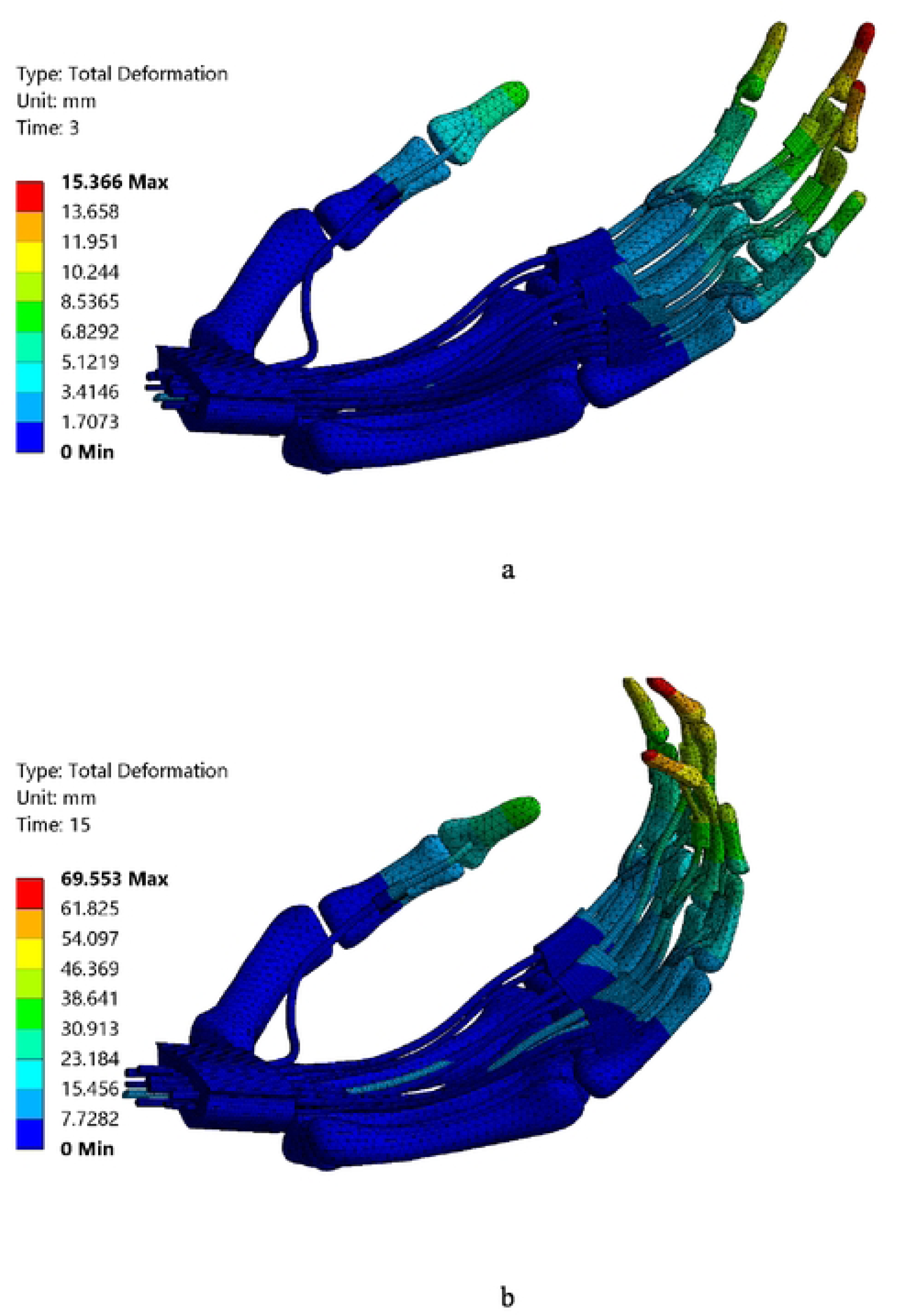
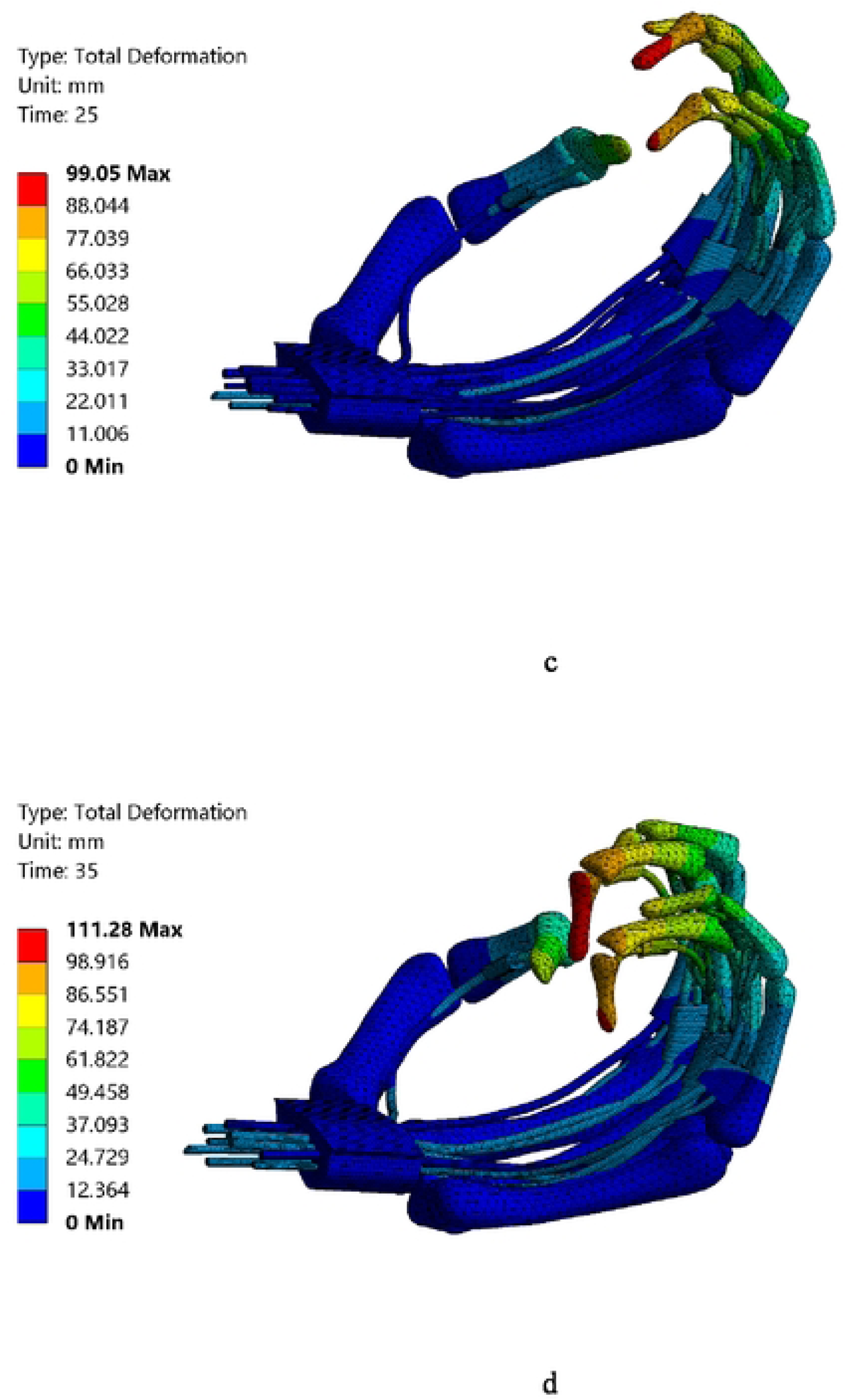
Total deformation during finger flexion: (a) *t* = 3s, (b) *t*=15s, (c), *t*= 25s, (d) *t*=35s.

**Table 4.** Maximum von Mises stress on the tendons during hand movements

| Movement type | Maximum Von Mises stress, Pa | Maximum Von Mises stress location |
| --- | --- | --- |
| Finger flexion | 4056 | Pinky deep |
| Wrist flexion (Bonded) | 15568 | Middle deep |
| Wrist flexion (No separation) | 8415 | Middle deep |
| Wrist extension (Bonded) | 16506 | Middle deep |
| Wrist extension (No separation) | 8530 | Thumb |

**Fig 6.**
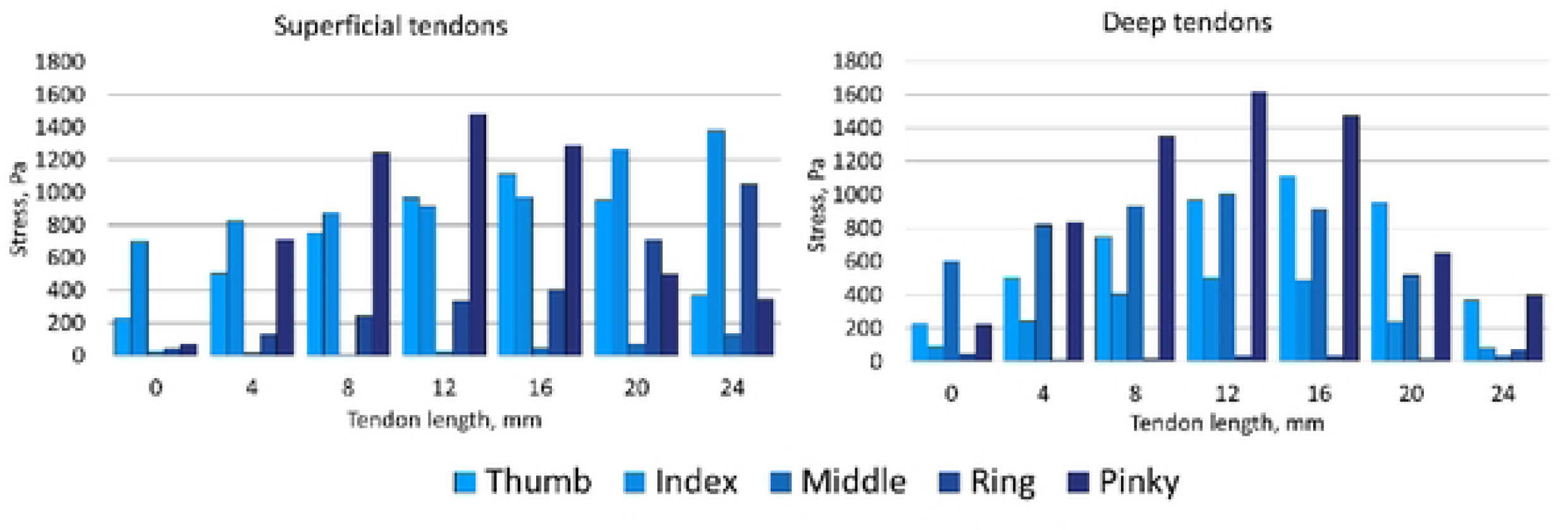
Tendons stress along tendon length (midline) during finger flexion: (a) superficial tendons, (b) deep tendons

### Wrist flexion and extension

Total deformation of tendons, median nerve, and connective tissue (hand is hide to focus on carpal tunnel) during flexion and extension of the hand with bonded contact is shown in (Fig 7, 8 at various modelling time *t*=1s, *t*=15s). Two types of tendons and connective tissue connections were considered during wrist flexion and extension. Stress along the midline of the tendon is shown in (Fig 9-12). Tendons were divided into two groups (deep and superficial), similar to the results for finger flexion. The thumb tendon is included in both groups as well. Results are shown for two types of contact for each tendon.

**Fig 7.**
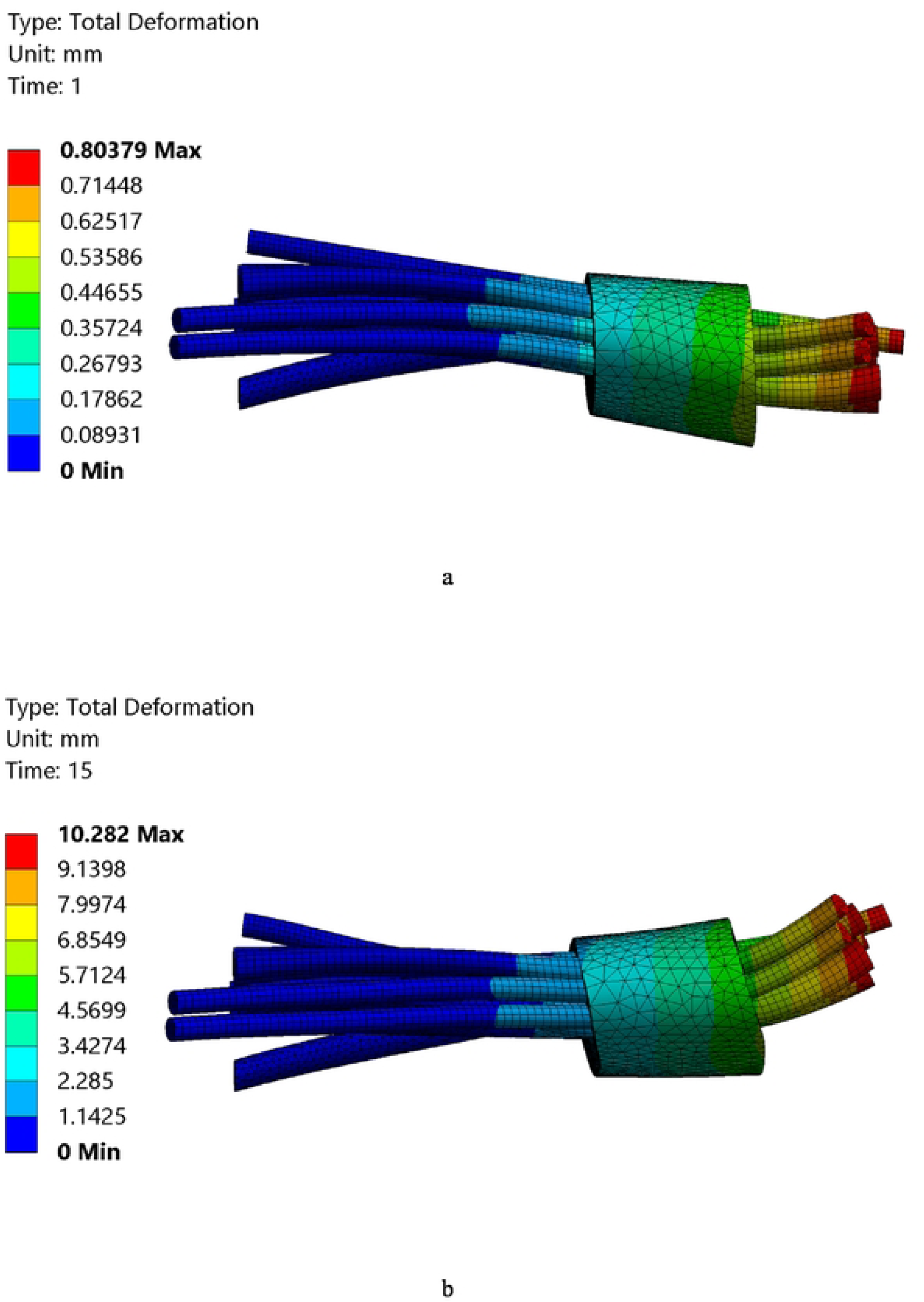
Total deformation during wrist flexion: (a) *t* = 1s, (b) *t*=15s.

**Fig 8.**
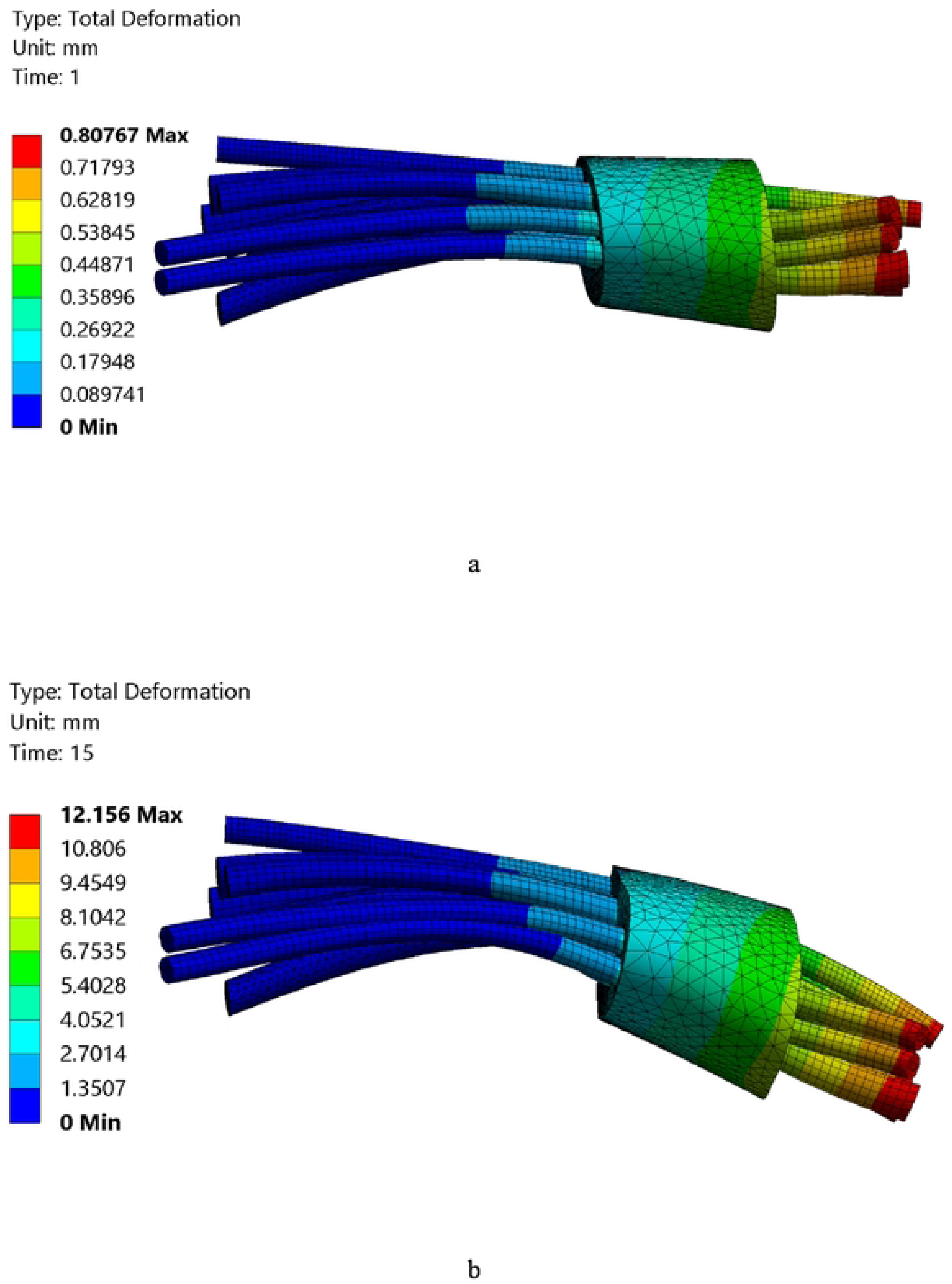
Total deformation during wrist extension: (a) *t* = 1s, (b) *t*=15s.

**Fig 9.**
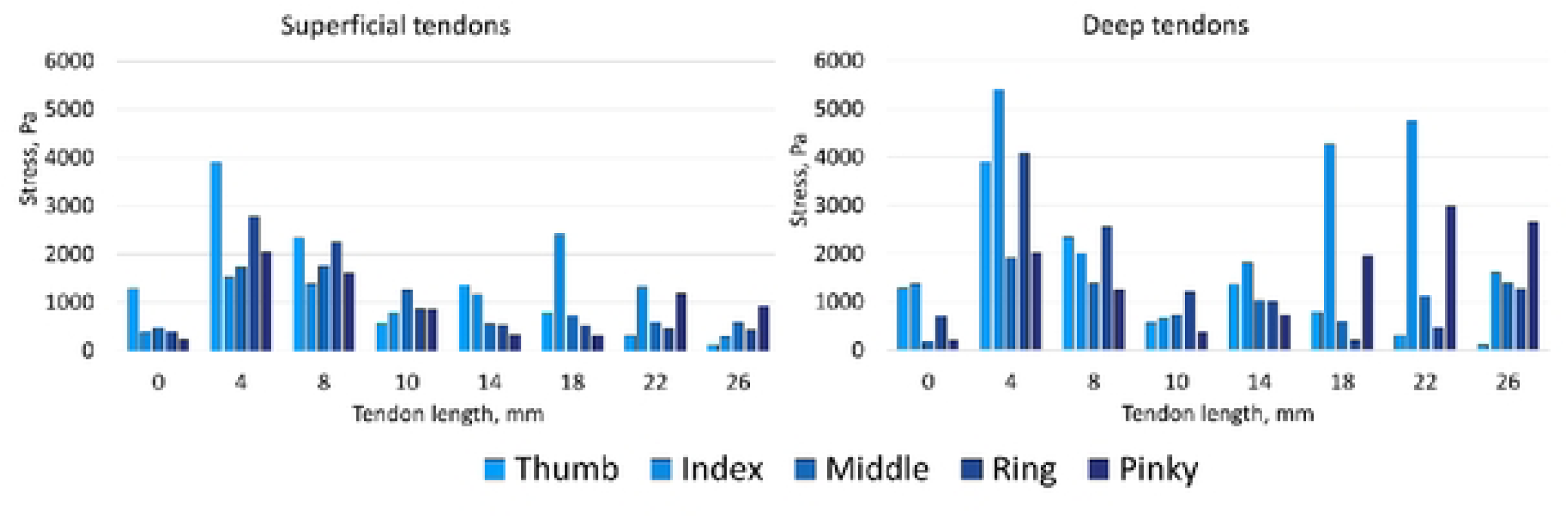
Stress in the tendons during wrist extension with bonded contact type along midline: (a) superficial tendons, (b) deep tendons.

**Fig 10.**
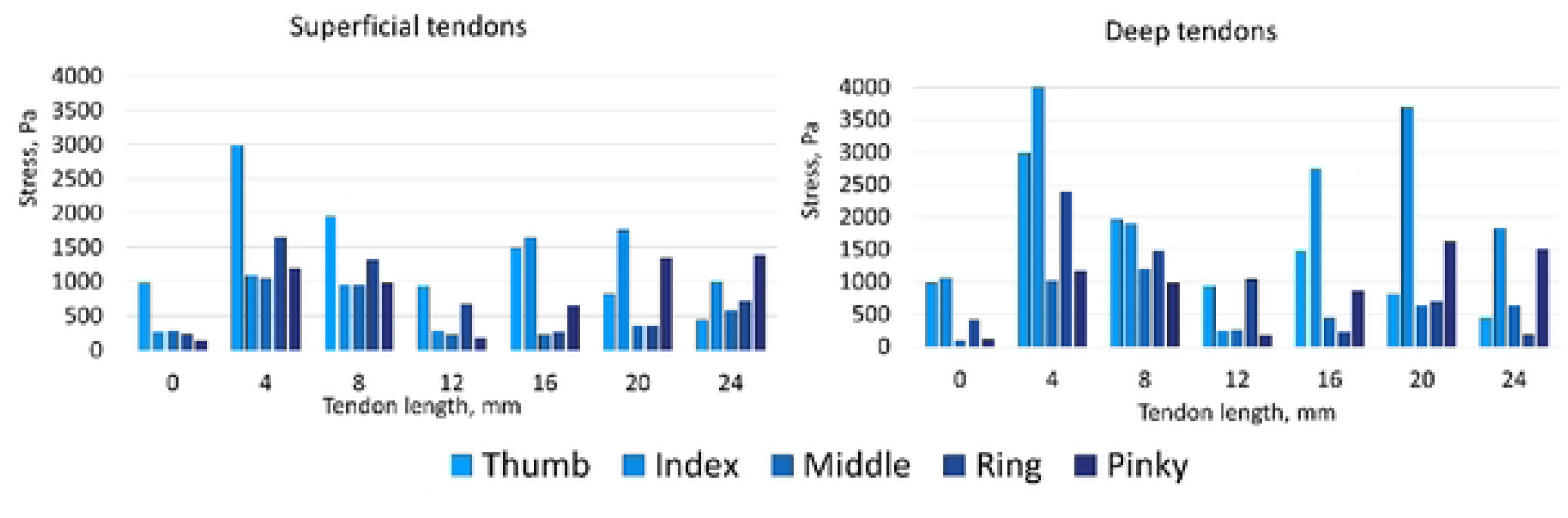
Stress in the tendons during wrist extension with no separation contact type along midline: (a) superficial tendons, (b) deep tendons.

**Fig 11.**
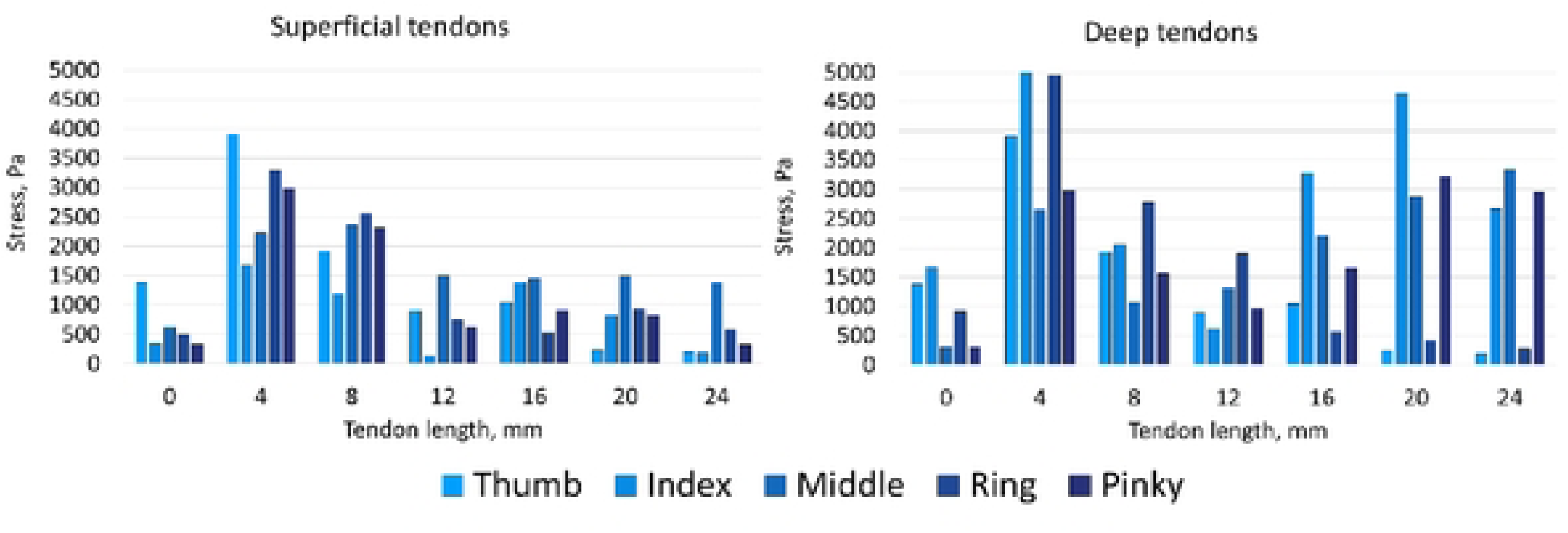
Stress in the tendons during wrist flexion with bonded contact type along midline: (a) superficial tendons, (b) deep tendons

**Fig 12.**
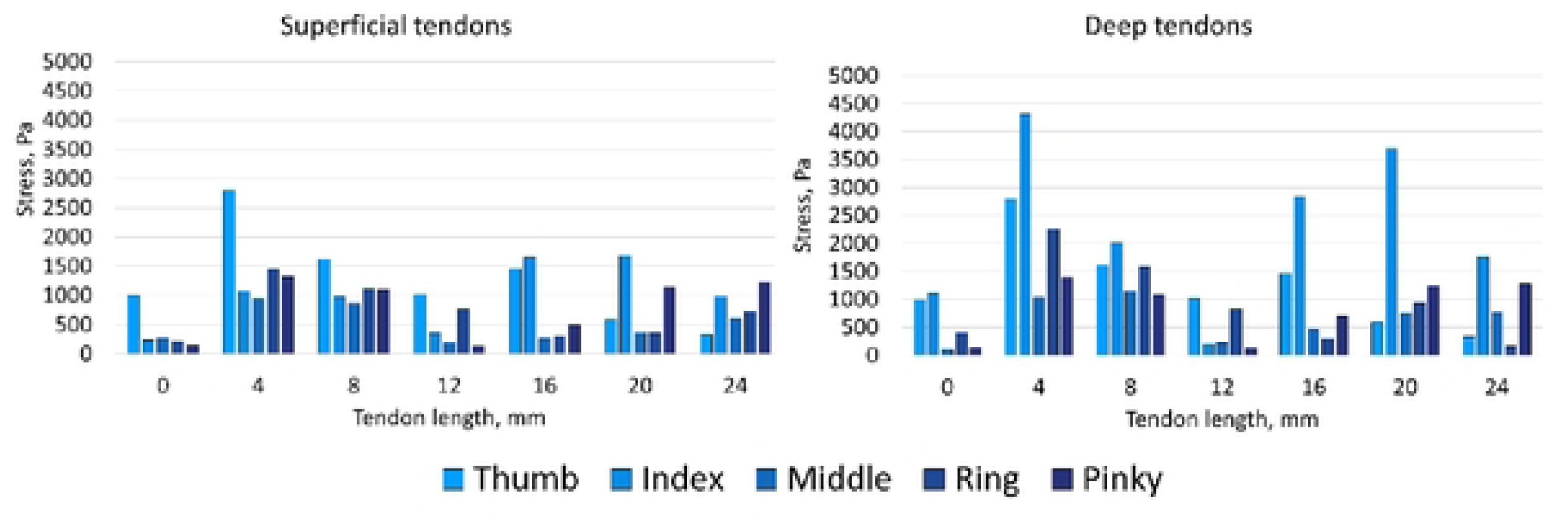
Stress in the tendons during wrist flexion with no separation contact type along midline: (a) superficial tendons, (b) deep tendons.

Finger joint flexion angles are usually determined by external factors. For example, in the movement of the shoulder or forearm, in various tissue injuries of the upper extremity or in neurophysiological disorders [73–75]. J.W. Lee and K.S.

Lee in their studies also used the motion capture method and the results were the same as the results of this study [76,77]. They revealed a non-linear character of fingers flexion angle plots, which was approved here also.

### Median nerve stress during finger flexion and wrist flexion/extension

Stress in the median nerve during finger flexion and during wrist flexion and extension was obtained according to the rotation angle of the respective phalanges. The human finger flexes in three joints: the joint between the distal phalanx and the middle phalanx (distal interphalangeal joint), the joint between the middle phalanx and the proximal phalanx (proximal interphalangeal joint) and the joint between the proximal phalanx and the metacarpal bones (metacarpal phalangeal joint). The angles at these joints are determined using motion capture in the third stage of this approach. To present the results, the maximum tension in the median nerve was presented as a function of the angle in the proximal interphalangeal joint. According to the results based on the hand motion capture, the angle in this joint changes faster than the others. The results of maximum median nerve stress during flexion and extension of the wrist are shown as a function of the angle of rotation at the wrist joint. Results presented for two types of connective tissue-tendon contact (Fig 13). Wrist flexion and extension was performed at 30 degrees, finger flexion was performed at 70 degrees. For convenience of comparison, the graph is shown up to 30 degrees. The median nerve stress changes further during finger flexion are close to linear.

**Fig 13.**
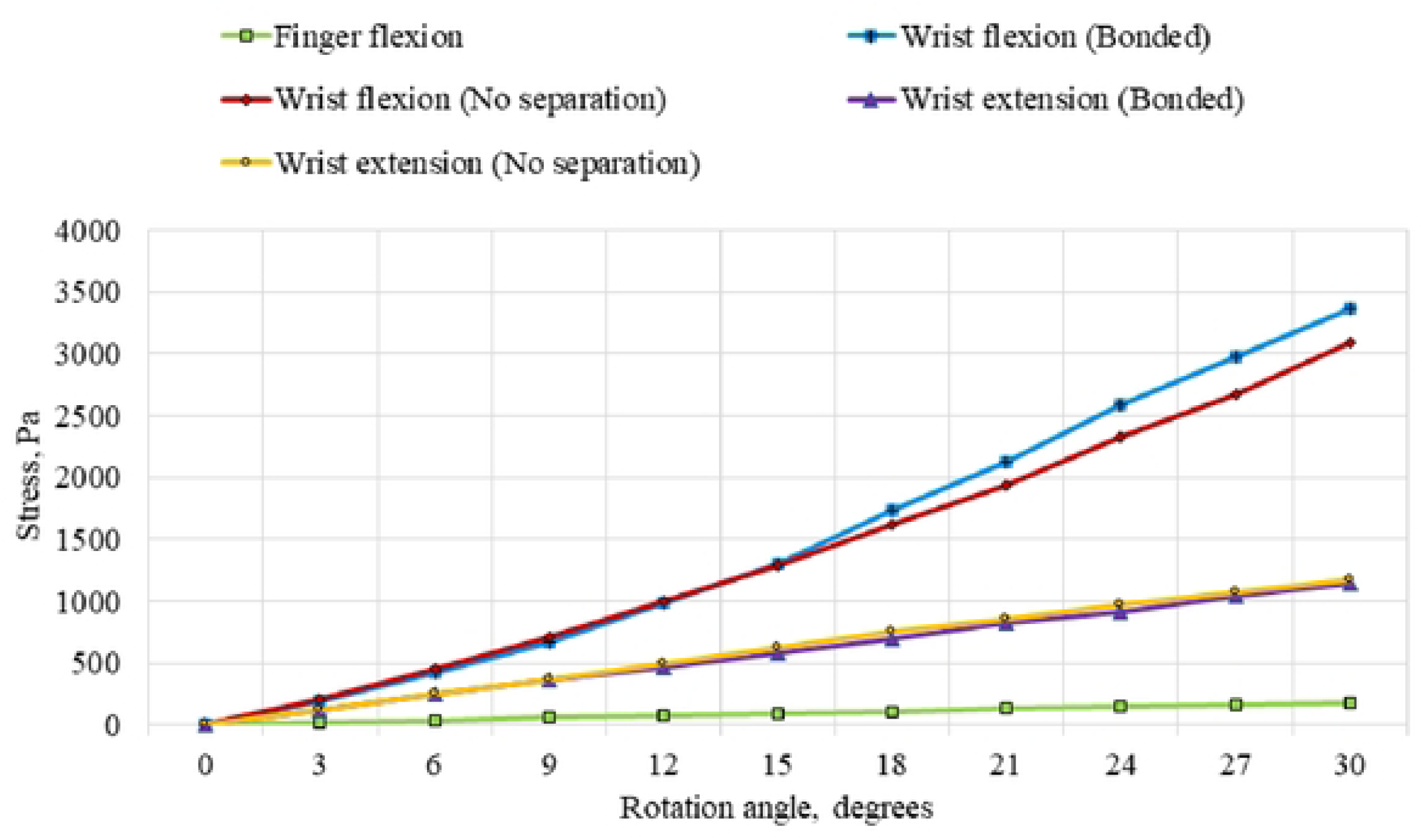
Relationship between maximum stresses in the median nerve and angle of rotation during finger flexion and wrist flexion/extension.

### Modelling of median nerve conduction during finger flexion and wrist flexion/extension

The shape change of the median nerve is accounted for by the function *r (z) = A-Bz-Cz^2^* By describing the shape of the median nerve during finger flexion and hand flexion and extension, coefficients *A, B* and *C* were obtained. The shape of the hand in flexion and extension appeared to be the same. The coefficient *C* is responsible for the non-linear curvature of the median nerve surface. In the finite element analysis, the shape of the median nerve was changed only linearly, so that coefficient *C* = 0 and coefficients *A* and *B* were 0.01 and 80 for finger flexion and 0.01 and 20 for hand flexion, respectively. The dependence of the membrane potential of the deformed median nerve on time at its various areas is shown at (Fig 14).

**Fig 14.**
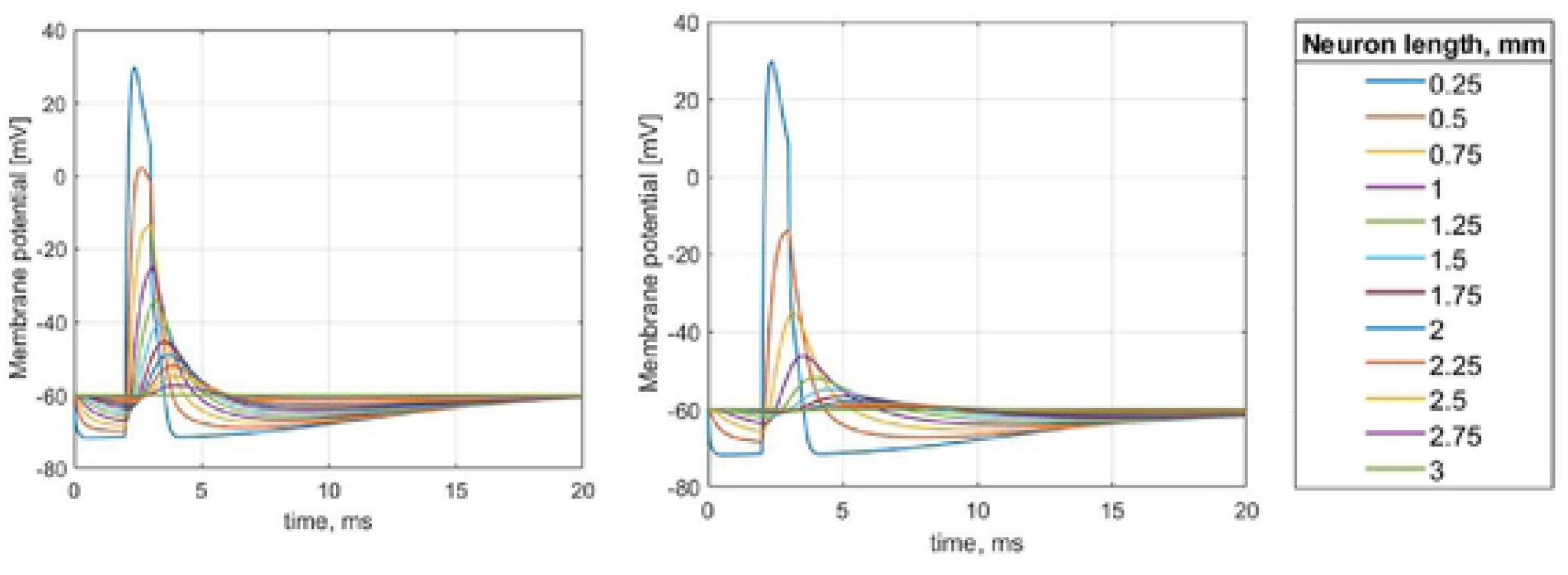
Dependence of the membrane potential of the deformed median nerve on time at its various areas: (a) – finger flexion (b) – wrist flexion.

## Discussion

The aim of this work was to create and apply a technique for early diagnosis of carpal tunnel syndrome. This study combines MRI / FEM / motion capture techniques to simulate carpal tunnel syndrome more precisely.

Stress was obtained in the median nerve and tendons during finger flexion and during flexion and extension of the hand. The loads in the carpal tunnel caused by finger flexion and extension were shown to be lower than those caused by hand flexion and extension [50,78,79]. The tendons slide along the median nerve and barely press on the nerve when fingers are flexed. However, when flexing and extending the hand, the tendons bend along the carpal tunnel and press on the median nerve [80]. Because the pinky tendons bend more than other tendons in the carpal canal due to a reduction in the cross-sectional area of the carpal tunnel, they experience higher stress during finger flexion than other tendons [81]. Fig 5 shows that the maximum stress values in the tendons are predominantly in the middle of the carpal tunnel. Low stress values in the tendons of the middle finger are related to the median nerve being located near them and the load is transferred to the nerve. The total tension of the deep tendons is 10% higher than the superficial tendons. This is due to the deep tendons of the fingers are more stretched than the superficial tendons [82,83].

Two contact types between tendons and connective tissue were considered during hand flexion and extension. The first type was an inseparable connection of tissues (bonded), the second type allowed the tissues to slide relative to each other, but at the same time without detaching (no separation). In this case, the results enable to analyze the difference in contact types and the difference in flexion and extension of the hand. The difference between hand flexion and extension with the bonded connection type was 5.74%. The difference between hand flexion and extension with the no separation connection type was 3.1%. It can be said that the tendon tension during hand flexion and extension is insufficient [84,85]. This can also be noted in the results of the median nerve tension of (Fig 14). The graph shows that the flexion of the hand and the extension differ slightly. However, the difference between the contact types was 31.7% for hand extension and 59.9% for hand flexion. This suggests a strong influence of the type of contact between the tendon and the connective tissue [86,87]. In clinical practice, an increase in carpal tunnel pressure may indicate an increase in tendon sliding friction coefficient along the carpal tunnel [88,89]. This manifestation can be a consequence of a lack of vitamins in the body or a metabolic disorder [90]. In almost all cases, the maximum stress was in the index finger tendons. This may be due to the increased size of the tendons caused by frequent movements of the patient’s index finger. The results obtained for stress in the tendons correspond to the values obtained in a recent paper by Lv. Y. et. al [43].

The maximum von Mises stress in the median nerve during flexion/extension of the hand are significantly higher than during finger flexion. When the fingers were flexed at 30 degrees, the median nerve stress was 177 Pa; when the hand was flexed and extended at 30 degrees with the Bonded contact type, it was 3364.3 and 3087 Pa, respectively; when the hand was flexed and extended at 30 degrees with the no separation contact type, it was 1143 and 1174 Pa, respectively. The loads acting on the median nerve were experimentally determined in the studies [91–93] and coincide with the results obtained in this article. In this method, the median nerve conduction depends on the electrophysiological parameters of the H-H model and the conduction equation, as well as on the shape of the median nerve described by the function *r(z).* Thus, conduction during finger flexion is attenuated along the length of the tendon slower than during flexion and extension of the hand [94,95]. This technique currently has no criteria for determining the presence or absence of carpal tunnel syndrome, because the method was tested on a completely healthy middle-aged woman who does not have carpal tunnel syndrome. Similar studies on a group of patients are needed to create an accurate criterion. However, this paper shows all the stages of the developed approach and shows the result of its work on a person.

The finite element method has many varying parameters that can greatly affect the obtained results. In that case, a comparison of results with experimental data is necessary. Gelberman et. al., Szabo et. al., Werner et. al., Rojviroj et. al., Hamanaka et. al. and Goss et. al. in their studies [91,96–101] presented results of experimental intracarpal pressure determination. Comparison of the results obtained in this study with the relevant experimental results from these works are presented in (Table 5). However, the experimental technique of pressure determination in the carpal tunnel allows to determine the fluid pressure within the carpal tunnel. The technique used in this study allows to determine the stress of all carpal tunnel tissues. In this case, the tendon and median nerve stress can be very different. Tendon stress correlates better with experimental data than median nerve stress. However, in some cases, a good correlation with median nerve tension was also shown. In this work, a healthy patient without carpal tunnel syndrome was examined according to the approach. Experimental studies most often focus on patients with carpal tunnel syndrome. Mouzakis et. al. and Lv et. al. in their studies [43,46] determined the stress in the tissues of the carpal tunnel also by the finite element method. Mouzakis et. et al. determined the stress of all tissues of the carpal tunnel during mouse and keyboard operation. Lv et. al. determined the stress in the finger flexor tendons.

**Table 5.**
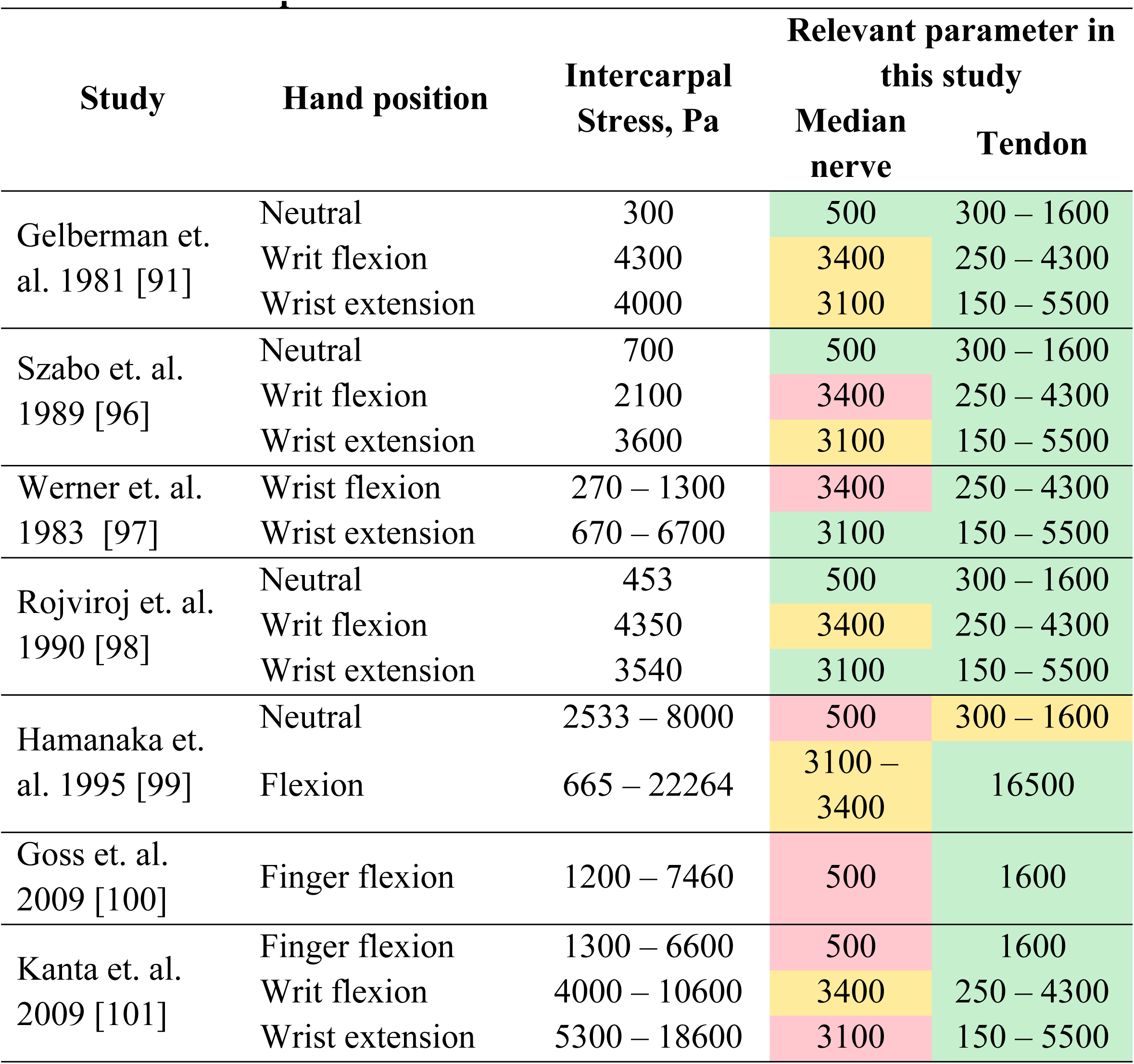
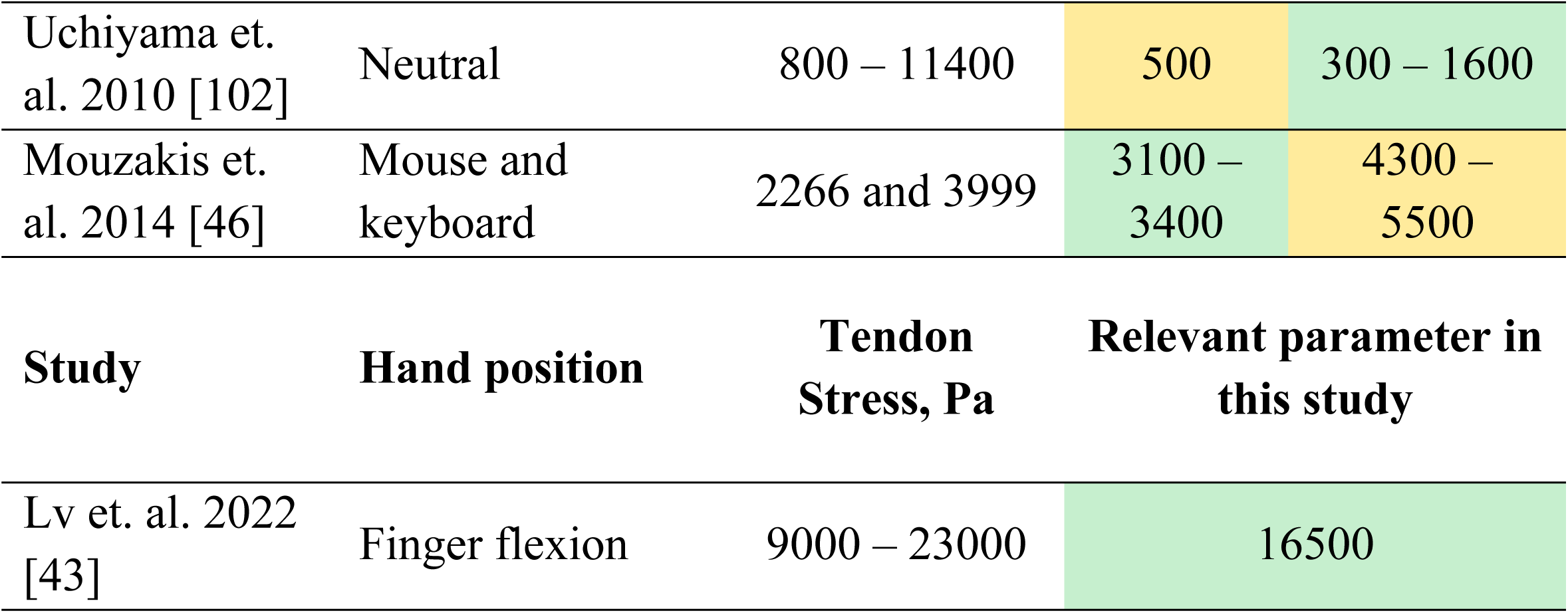
Comparison of results with other studies

## Limitations

The construction of patient–specific geometry is associated with subjective factors. The choice of the MRI method of operation and the processing of the resulting geometry may lead to minor changes in the calculation results. Nevertheless, in this paper, the construction of an ideal personal geometry may be a limitation.

When using the FEM, it is necessary to set the mechanical parameters of the studied tissues. In this paper, these parameters were taken from the experimental work of other authors. Although these parameters may be different for different people, and may also change during life. Determining these parameters in a live person is difficult at the moment. The choice of mechanical parameters of the simulated tissues is a limitation of the proposed technique. Nevertheless, these parameters can be determined experimentally in cadavers and the results obtained can be divided into groups. For example, the parameters that characterize men and women, and also divide these groups by age. This may be a further line of research. The rotation axis location during hand flexion and extension in this study was made on the geometry of the carpal tunnel. Although, during hand flexion, the connected to each other by ligaments carpal bones move. Bone–ligamentous complex determines the axis of rotation of the hand. In this case, the location of the axis of rotation may change. Determining the rotation axis during flexion and extension of the hand can also be a direction of further research.

## Conclusions

This study presents a six–stage combined approach for the early diagnosis of carpal tunnel syndrome. Each stage aims to improve understanding of the mechanical and neurophysiological factors influencing the development of carpal tunnel syndrome. The finite element method is used to determine the mechanical stress in the carpal tunnel. MRI and CT scans of the patient’s hand were used to create the patient–specific geometry. Software has been created to capture hand motion and is used to determine the personal movement of the patient’s fingers and hand. The obtained stress–strain state of the median nerve is used to model the conductivity of the electrical impulse along the median nerve. The Hodgkin and Huxley model and the extended cable equation describe nerve conduction in this approach. The results showed that compression of the median nerve is highly affected by the contact condition of the tendon and connective tissue. It has been shown that hand flexion reduces conduction in the median nerve more than finger flexion. The developed approach can be introduced into medical practice to improve the quality for early diagnostics of the carpal tunnel syndrome.

## Data Availability

-

## Acknowledgements

This work was supported by Russian Science Foundation and Perm Region (No. 22–21–20067, https://rscf.ru/en/project/22-21-20067).

## References

1. Núñez-Cortés R, Cruz-Montecinos C, Torreblanca-Vargas S, Andersen LL, Tapia C, Ortega-Palavecinos M, et al. Social determinants of health and physical activity are related to pain intensity and mental health in patients with carpal tunnel syndrome. Musculoskelet Sci Pract. 2023;63. doi:10.1016/j.msksp.2023.102723

2. Bickel KD. Carpal Tunnel Syndrome. J Hand Surg Am. 2010;35: 147–152. doi:10.1016/j.jhsa.2009.11.003

3. Sousa RL, Moraes VY de, Zobiole AF, Nakachima LR, Belloti JC. Diagnostic criteria and outcome measures in randomized clinical trials on carpal tunnel syndrome: a systematic review. Sao Paulo Med J. 2023;141: e2022086. doi:10.1590/1516-3180.2022.0086.07022023

4. Nazari G, Shah N, MacDermid JC, Woodhouse L. The Impact of Sensory, Motor and Pain Impairments on Patient-Reported and Performance Based Function in Carpal Tunnel Syndrome. Open Orthop J. 2017;11. doi:10.2174/1874325001711011258

5. Wipperman J, Goerl K. Diagnosis and management of carpal tunnel syndrome. J Musculoskelet Med. 2016;94: 47–60.

6. Nora DB, Becker J, Ehlers JA, Gomes I. What symptoms are truly caused by median nerve compression in carpal tunnel syndrome? Clin Neurophysiol. 2005;116. doi:10.1016/j.clinph.2004.08.013

7. Nunez F, Vranceanu AM, Ring D. Determinants of pain in patients with carpal tunnel syndrome. Clin Orthop Relat Res. 2010;468: 3328–3332. doi:10.1007/s11999-010-1551-x

8. Zuniga AF, Keir PJ. Diagnostic and research techniques in carpal tunnel syndrome. Crit Rev Biomed Eng. 2019;47. doi:10.1615/CritRevBiomedEng.2020030827

9. Dabbagh A, Ziebart C, MacDermid JC. Accuracy of diagnostic clinical tests and questionnaires in screening for carpal tunnel syndrome among workers-A systematic review. J Hand Ther. 2021;34. doi:10.1016/j.jht.2021.04.003

10. Elseddik M, Mostafa RR, Elashry A, El-Rashidy N, El-Sappagh S, Elgamal S, et al. Predicting CTS Diagnosis and Prognosis Based on Machine Learning Techniques. Diagnostics. 2023;13: 1–18. doi:10.3390/diagnostics13030492

11. Drăghici NC, Leucuța DC, Ciobanu DM, Stan AD, Lupescu TD, Mureșanu DF. Clinical Utility of Boston-CTS and Six-Item CTS Questionnaires in Carpal Tunnel Syndrome Associated with Diabetic Polyneuropathy. Diagnostics. 2023;13. doi:10.3390/diagnostics13010004

12. Levine DW, Simmons BP, Koris MJ, Daltroy LH, Hohl GG, Fossel AH, et al. A self-administered questionnaire for the assessment of severity of symptoms and functional status in carpal tunnel syndrome. J Bone Jt Surg. 1993;75. doi:10.2106/00004623-199311000-00002

13. Greenslade JR, Mehta RL, Belward P, Warwick DJ. Dash and boston questionnaire assessment of carpal tunnel syndrome outcome: What is the responsiveness of an outcome questionnaire? J Hand Surg Am. 2004;29 B. doi:10.1016/j.jhsb.2003.10.010

14. Chung KC, Hamill JB, Walters MR, Hayward RA. The Michigan hand outcomes questionnaire (MHQ): Assessment of responsiveness to clinical change. Ann Plast Surg. 1999;42. doi:10.1097/00000637-199906000-00006

15. Pransky G, Feuerstein M, Himmelstein J, Katz JN, Vickers-Lahti M. Measuring functional outcomes in work-related upper extremity disorders: Development and validation of the upper extremity function scale. J Occup Environ Med. 1997;39. doi:10.1097/00043764-199712000-00014

16. Leite JCDC, Jerosch-Herold C, Song F. A systematic review of the psychometric properties of the Boston Carpal Tunnel Questionnaire. BMC Musculoskeletal Disorders. 2006. doi:10.1186/1471-2474-7-78

17. Franchignoni F, Vercelli S, Giordano A, Sartorio F, Bravini E, Ferriero G. Minimal clinically important difference of the disabilities of the arm, shoulder and hand outcome measure (DASH) and its shortened version (quickDASH). J Orthop Sports Phys Ther. 2014;44. doi:10.2519/jospt.2014.4893

18. Crisp AH, Jones MG, Slater P. The Middlesex Hospital Questionnaire: A validity study. Br J Med Psychol. 1978;51. doi:10.1111/j.2044-8341.1978.tb02472.x

19. Ito M, Bentley KH, Oe Y, Nakajima S, Fujisato H, Kato N, et al. Assessing depression related severity and functional impairment(warning) the Overall Depression Severity and Impairment Scale (ODSIS). PLoS One. 2015;10. doi:10.1371/journal.pone.0122969

20. Sambandam SN, Priyanka P, Gul A, Ilango B. Critical analysis of outcome measures used in the assessment of carpal tunnel syndrome. Int Orthop. 2008;32. doi:10.1007/s00264-007-0344-7

21. Zhang D, Chruscielski CM, Blazar P, Earp BE. Accuracy of Provocative Tests for Carpal Tunnel Syndrome. J Hand Surg Glob Online. 2020;2. doi:10.1016/j.jhsg.2020.03.002

22. Dabbagh A, MacDermid JC, Yong J, Packham TL, Macedo LG, Ghodrati M. Diagnostic accuracy of sensory and motor tests for the diagnosis of carpal tunnel syndrome: a systematic review. BMC Musculoskelet Disord. 2021;22. doi:10.1186/s12891-021-04202-y

23. Agarwal V, Singh R, Sachdev A, Wiclaff, Shekhar S, Goel D. A prospective study of the long-term efficacy of local methyl prednisolone acetate injection in the management of mild carpal tunnel syndrome. Rheumatology. 2005;44. doi:10.1093/rheumatology/keh571

24. Liu D, Ahmet A, Ward L, Krishnamoorthy P, Mandelcorn ED, Leigh R, et al. A practical guide to the monitoring and management of the complications of systemic corticosteroid therapy. Allergy, Asthma and Clinical Immunology. 2013. doi:10.1186/1710-1492-9-30

25. Osiak K, Mazurek A, Pękala P, Koziej M, Walocha JA, Pasternak A. Electrodiagnostic studies in the surgical treatment of carpal tunnel syndrome—a systematic review. Journal of Clinical Medicine. 2021. doi:10.3390/jcm10122691

26. Jablecki CK, Andary MT, Floeter MK, Miller RG, Quartly CA, Vennix MJ, et al. Practice parameter: Electrodiagnostic studies in carpal tunnel syndrome: Report of the American Association of Electrodiagnostic Medicine, American Academy of Neurology, and the American Academy of Physical Medicine and Rehabilitation. Neurology. 2002;58. doi:10.1212/WNL.58.11.1589

27. Witt JC, Hentz JG, Stevens JC. Carpal tunnel syndrome with normal nerve conduction studies. Muscle and Nerve. 2004;29. doi:10.1002/mus.20019

28. Park JS, Won HC, Oh JY, Kim DH, Hwang SC, Yoo J Il. Value of cross-sectional area of median nerve by MRI in carpal tunnel syndrome. Asian J Surg. 2020;43. doi:10.1016/j.asjsur.2019.08.001

29. McDonagh C, Alexander M, Kane D. The role of ultrasound in the diagnosis and management of carpal tunnel syndrome: A new paradigm. Rheumatol (United Kingdom). 2014;54: 9–19. doi:10.1093/rheumatology/keu275

30. Lam KHS, Wu Y-T, Reeves KD, Galluccio F, Allam AE-S, Peng PWH. Ultrasound-Guided Interventions for Carpal Tunnel Syndrome: A Systematic Review and Meta-Analyses. Diagnostics. 2023;13. doi:10.3390/diagnostics13061138

31. Atroshi I, Gummesson C, Johnsson R, Ornstein E, Ranstam J, Rosen I. [Prevalence for clinically proved carpal tunnel syndrome is 4 percent]. Lakartidningen. 2000;97.

32. Musculoskeletal Ultrasonography in Rheumatic Diseases. Musculoskeletal Ultrasonography in Rheumatic Diseases. 2015. doi:10.1007/978-3-319-15723-8

33. Byra M, Hentzen E, Du J, Andre M, Chang EY, Shah S. Assessing the Performance of Morphologic and Echogenic Features in Median Nerve Ultrasound for Carpal Tunnel Syndrome Diagnosis. J Ultrasound Med. 2020;39. doi:10.1002/jum.15201

34. Kuo TT, Lee MR, Liao YY, Chen JP, Hsu YW, Yeh CK. Assessment of median nerve mobility by ultrasound dynamic imaging for diagnosing carpal tunnel syndrome. PLoS One. 2016;11. doi:10.1371/journal.pone.0147051

35. Kanagasabai K. Ultrasound of Median Nerve in the Diagnosis of Carpal Tunnel Syndrome-Correlation with Electrophysiological Studies. Indian J Radiol Imaging. 2022;32. doi:10.1055/s-0041-1741088

36. Cebral JR, Lhner R. From medical images to anatomically accurate finite element grids. Int J Numer Methods Eng. 2001;51. doi:10.1002/nme.205

37. Peshin SE, Karakulova YV, Nyashin YI, Nyashin MM. Carpal tunnel syndrome in terms of biomechanics. Literature review. Russ J Biomech. 2022;26: 9–13. Available: https://vestnik.pstu.ru/biomech/archives/?id=&folder_id=10825

38. Ko C, Brown TD. A fluid-immersed multi-body contact finite element formulation for median nerve stress in the carpal tunnel. Comput Methods Biomech Biomed Engin. 2007;10. doi:10.1080/10255840701430480

39. Guo X, Fan Y, Li ZM. Three dimensional finite element analysis on the morphological change of the transverse carpal ligament. 2007 IEEE/ICME International Conference on Complex Medical Engineering, CME 2007. 2007. doi:10.1109/ICCME.2007.4382071

40. Wei Y, Zou Z, Wei G, Ren L, Qian Z. Subject-Specific Finite Element Modelling of the Human Hand Complex: Muscle-Driven Simulations and Experimental Validation. Ann Biomed Eng. 2020;48. doi:10.1007/s10439-019-02439-2

41. Perevoshchikova N, Moerman KM, Akhbari B, Bindra R, Maharaj JN, Lloyd DG, et al. Finite element analysis of the performance of additively manufactured scaffolds for scapholunate ligament reconstruction. PLoS One. 2021;16. doi:10.1371/journal.pone.0256528

42. Henderson J, Thoreson A, Yoshii Y, Zhao KD, Amadio PC, An KN. Finite element model of subsynovial connective tissue deformation due to tendon excursion in the human carpal tunnel. J Biomech. 2011;44. doi:10.1016/j.jbiomech.2010.09.001

43. Lv Y, Zheng Q, Chen X, Hou C, An M. Analysis on synergistic cocontraction of extrinsic finger flexors and extensors during flexion movements: A finite element digital human hand model. PLoS One. 2022;17: 1–15. doi:10.1371/journal.pone.0268137

44. Yao Y, Erdemir A, Li ZM. Finite element analysis for transverse carpal ligament tensile strain and carpal arch area. J Biomech. 2018;73. doi:10.1016/j.jbiomech.2018.04.005

45. Chang CT, Chen YH, Lin CCK, Ju MS. Finite element modeling of hyper-viscoelasticity of peripheral nerve ultrastructures. J Biomech. 2015;48. doi:10.1016/j.jbiomech.2015.04.004

46. Mouzakis DE, Rachiotis G, Zaoutsos S, Eleftheriou A, Malizos KN. Finite element simulation of the mechanical impact of computer work on the carpal tunnel syndrome. J Biomech. 2014;47. doi:10.1016/j.jbiomech.2014.07.004

47. Javanmardian A, HaghPanahi M. 3 dimensional finite element analysis of the human wrist joint without ligaments under compressive loads. 2010 17th Iranian Conference of Biomedical Engineering, ICBME 2010 - Proceedings. 2010. doi:10.1109/ICBME.2010.5704996

48. Oflaz H, Gunal I. Maximum loading of carpal bones during movements: a finite element study. Eur J Orthop Surg Traumatol. 2019;29. doi:10.1007/s00590-018-2287-7

49. Yokota H, Yasui M, Hirai S, Hatayama N, Ohshima S, Nakano T, et al. Evaluation of the pressure on the dorsal surface of the distal radius using a cadaveric and computational model: clinical considerations in intersection syndrome and Colles’ fracture. Anat Sci Int. 2020;95. doi:10.1007/s12565-019-00491-5

50. Peshin S, Karakulova Y, Kuchumov AG. Finite Element Modeling of the Fingers and Wrist Flexion/Extension Effect on Median Nerve Compression. Appl Sci. 2023;13. doi:10.3390/app13021219

51. Osamura N, Zhao C, Zobitz ME, An KN, Amadio PC. Evaluation of the material properties of the subsynovial connective tissue in carpal tunnel syndrome. Clin Biomech. 2007;22. doi:10.1016/j.clinbiomech.2007.07.009

52. Matsuura Y, Thoreson AR, Zhao C, Amadio PC, An KN. Development of a hyperelastic material model of subsynovial connective tissue using finite element modeling. J Biomech. 2016;49. doi:10.1016/j.jbiomech.2015.09.048

53. Main EK, Goetz JE, Baer TE, Klocke NF, Brown TD. Volar/dorsal compressive mechanical behavior of the transverse carpal ligament. J Biomech. 2012;45. doi:10.1016/j.jbiomech.2012.01.048

54. Main EK, Goetz JE, James Rudert M, Goreham-Voss CM, Brown TD. Apparent transverse compressive material properties of the digital flexor tendons and the median nerve in the carpal tunnel. J Biomech. 2011;44. doi:10.1016/j.jbiomech.2010.12.005

55. Ogden R. Non-linear elastic deformations. Eng Anal Bound Elem. 1984;1. doi:10.1016/0955-7997(84)90049-3

56. Lundborg G, Gelberman RH, Minteer-Convery M, Lee YF, Hargens AR. Median nerve compression in the carpal tunnel—Functional response to experimentally induced controlled pressure. J Hand Surg Am. 1982;7. doi:10.1016/S0363-5023(82)80175-5

57. Rempel D, Dahlin L, Lundborg G. Pathophysiology of nerve compression syndromes: Response of peripheral nerves to loading. Journal of Bone and Joint Surgery. 1999. doi:10.2106/00004623-199911000-00013

58. Topp KS, Boyd BS. Structure and biomechanics of peripheral nerves: Nerve responses to physical stresses and implications for physical therapist practice. Physical Therapy. 2006. doi:10.1093/ptj/86.1.92

59. Mittal P, Shenoy S, Sandhu JS. Effect of different cuff widths on the motor nerve conduction of the median nerve: An experimental study. J Orthop Surg Res. 2008;3. doi:10.1186/1749-799X-3-1

60. Hodgkin A, Huxley A. A quantitative description of membrane current and its application to conductance and excitation. J Physiol. 1952;117: 500–44.

61. FitzHugh R. Impulses and Physiological States in Theoretical Models of Nerve Membrane. Biophys J. 1961;1. doi:10.1016/S0006-3495(61)86902-6

62. Plonsey R, Barr RC. Bioelectricity: A quantitative approach. Bioelectricity: A Quantitative Approach. 2007. doi:10.1007/978-0-387-48865-3

63. Jérusalem A, García-Grajales JA, Merchán-Pérez A, Peña JM. A computational model coupling mechanics and electrophysiology in spinal cord injury. Biomech Model Mechanobiol. 2014;13. doi:10.1007/s10237-013-0543-7

64. Tekieh T, Shahzadi S, Rafii-Tabar H, Sasanpour P. Are deformed neurons electrophysiologically altered? A simulation study. Curr Appl Phys. 2016;16: 1413–1417. doi:10.1016/j.cap.2016.07.012

65. Cavarretta F, Naldi G. Mathematical study of a nonlinear neuron model with active dendrites. AIMS Math. 2019;4. doi:10.3934/math.2019.3.831

66. Tian J, Huang G, Lin M, Qiu J, Sha B, Lu TJ, et al. A mechanoelectrical coupling model of neurons under stretching. J Mech Behav Biomed Mater. 2019;93. doi:10.1016/j.jmbbm.2019.02.007

67. Wu YT, Gilpin K, Adnan A. Effects of Focal Axonal Swelling Level on the Action Potential Signal Transmission. J Comput Neurosci. 2020;48. doi:10.1007/s10827-020-00750-9

68. Nazari-Vanani R, Mohammadpour R, Asadian E, Rafii-Tabar H, Sasanpour P. A computational modelling study of excitation of neuronal cells with triboelectric nanogenerators. Sci Rep. 2022;12: 1–10. doi:10.1038/s41598-022-17050-0

69. Bazarevsky V, Zhang F. On-Device, Real-Time Hand Tracking with MediaPipe. In: Google AI Blog. 2019.

70. Pistoia W, Van Rietbergen B, Lochmüller EM, Lill CA, Eckstein F, Rüegsegger P. Estimation of distal radius failure load with micro-finite element analysis models based on three-dimensional peripheral quantitative computed tomography images. Bone. 2002;30. doi:10.1016/S8756-3282(02)00736-6

71. Ma Z, Hu S, Tan JS, Myer C, Njus NM, Xia Z. In vitro and in vivo mechanical properties of human ulnar and median nerves. J Biomed Mater Res - Part A. 2013;101 A. doi:10.1002/jbm.a.34573

72. Armstrong J, Chaffins DONB. Some Biomechanical Aspects of the carpal tunnel. Biomechanics. 1979;12: 567–570.

73. Tan SR, Mathis LM, El-Gamal HM. Surgical Pearl: Safe splinting positions for skin grafts on the hand and wrist. J Am Acad Dermatol. 2005;52. doi:10.1016/j.jaad.2004.10.866

74. Taams KO, Ash GJ, Johannes S. Maintaining the safe position in a palmar splint: The “double-T” plaster splint. J Hand Surg (British Eur Vol. 1996;21. doi:10.1016/S0266-7681(05)80214-1

75. Clark DC. Common Acute Hand Infections. American Family Physician. 2003.

76. Lee KS, Jung MC. Flexion and extension angles of resting fingers and wrist. Int J Occup Saf Ergon. 2014;20. doi:10.1080/10803548.2014.11077038

77. Lee JW, Rim K. Measurement of finger joint angles and maximum finger forces during cylinder grip activity. J Biomed Eng. 1991;13: 152–162. doi:10.1016/0141-5425(91)90062-C

78. Li ZM, Jordan DB. Carpal tunnel mechanics and its relevance to carpal tunnel syndrome. Hum Mov Sci. 2023;87. doi:10.1016/j.humov.2022.103044

79. Aletto C, Aicale R, Oliva F, Maffulli N. Hand Flexor Tendon Repair: From Biology to Surgery and Rehabilitation. Hand Clin. 2023;39: 215–225. doi:10.1016/j.hcl.2022.12.001

80. Toge Y, Nishimura Y, Basford JR, Nogawa T, Yamanaka M, Nakamura T, et al. Comparison of the effects of flexion and extension of the thumb and fingers on the position and cross-sectional area of the median nerve. PLoS One. 2013;8. doi:10.1371/journal.pone.0083565

81. Murthy PG, Bae DS. Injuries to the Wrist, Hand, and Fingers. 2018. doi:10.1007/978-3-319-56188-2_10

82. Portenard AC, Pegot A, Lievain L, Michelin P, Angot É, Beccari R, et al. The distal dorsal intermetacarpal ligament: characterization of an overlooked structure—an anatomical study of 25 hands. Surg Radiol Anat. 2023;45: 673– 679. doi:10.1007/s00276-023-03139-2

83. Gardenier J, Garg R, Mudgal C. Upper extremity tendon transfers: A brief review of history, common applications, and technical tips. Indian Journal of Plastic Surgery. 2020. doi:10.1055/s-0040-1716456

84. Abdelhafiz MH, Andreasen Struijk LNS, Dosen S, Spaich EG. Biomimetic Tendon-Based Mechanism for Finger Flexion and Extension in a Soft Hand Exoskeleton: Design and Experimental Assessment. Sensors. 2023;23. doi:10.3390/s23042272

85. Kim J, Rhee SH, Gong HS, Oh S, Baek GH. Biomechanical analyses of the human flexor tendon adhesion models in the hand: A cadaveric study. J Orthop Res. 2015;33. doi:10.1002/jor.22798

86. Mackey AL, Heinemeier KM, Koskinen SOA, Kjaer M. Dynamic adaptation of tendon and muscle connective tissue to mechanical loading. Connect Tissue Res. 2008;49. doi:10.1080/03008200802151672

87. Turrina A, Martínez-González MA, Stecco C. The muscular force transmission system: Role of the intramuscular connective tissue. J Bodyw Mov Ther. 2013;17. doi:10.1016/j.jbmt.2012.06.001

88. Ettema AM, Zhao C, Amadio PC, O’Byrne MM, An KN. Gliding characteristics of flexor tendon and tenosynovium in carpal tunnel syndrome: A pilot study. Clin Anat. 2007;20. doi:10.1002/ca.20379

89. Farias Zuniga A, Keir PJ. Thirty Minutes of Sub-diastolic Blood Flow Occlusion Alters Carpal Tunnel Tissue Function and Mechanics. Ultrasound Med Biol. 2022;48. doi:10.1016/j.ultrasmedbio.2022.02.008

90. Waugh CM, Mousavizadeh R, Lee J, Screen HRC, Scott A. Mild hypercholesterolemia impacts achilles sub-tendon mechanical properties in young rats. BMC Musculoskelet Disord. 2023;24: 282. doi:10.1186/s12891-023-06375-0

91. Bauman TD, Gelberman RH, Mubarak SJ, Garfin SR. The acute carpal tunnel syndrome. Clin Orthop Relat Res. 1981;156: 151–156. doi:10.1097/00003086-198105000-00019

92. Lundborg G, Myers R, Powell H. Nerve compression injury and increased endoneurial fluid pressure: A quot;miniature compartment syndromequot; J Neurol Neurosurg Psychiatry. 1983;46. doi:10.1136/jnnp.46.12.1119

93. Luchetti R, Schoenhuber R, Alfarano M, Deluca S, De Cicco G, Landi A. Carpal tunnel syndrome: Correlations between pressure measurement and intraoperative electrophysiological nerve study. Muscle Nerve. 1990;13. doi:10.1002/mus.880131211

94. Min J, Choi T, Cha Y. Multiple Tendon-inspired Sensors for Hand Motion Detection. Smart Mater Struct. 2023;32. doi:10.1088/1361-665X/acafb9

95. Kursa K, Lattanza L, Diao E, Rempel D. In vivo flexor tendon forces increase with finger and wrist flexion during active finger flexion and extension. J Orthop Res. 2006;24. doi:10.1002/jor.20110

96. Szabo RM, Chidgey LK. Stress carpal tunnel pressures in patients with carpal tunnel syndrome and normal patients. J Hand Surg Am. 1989;14. doi:10.1016/0363-5023(89)90178-0

97. Werner CO, Elmqvist D, Ohlin P. Pressure and nerve lesion in the carpal tunnel. Acta Orthop. 1983;54. doi:10.3109/17453678308996576

98. Rojviroj S, Sirichativapee W, Kowsuwon W, Wongwiwattananon J, Tamnanthong N, Jeeravipoolvarn P. Pressures in the carpal tunnel. A comparison between patients with carpal tunnel syndrome and normal subjects. J Bone Jt Surg - Ser B. 1990;72. doi:10.1302/0301-620x.72b3.2187880

99. Hamanaka I, Okutsu I, Shimizu K, Takatori Y, Ninomiya S. Evaluation of carpal canal pressure in carpal tunnel syndrome. J Hand Surg Am. 1995;20. doi:10.1016/S0363-5023(05)80442-3

100. Goss BC, Agee JM. Dynamics of Intracarpal Tunnel Pressure in Patients With Carpal Tunnel Syndrome. J Hand Surg Am. 2010;35. doi:10.1016/j.jhsa.2009.09.019

101. Kanta M, Ehler E, Kremlácek J, Rehák S, Lastovicka D, Adamkov J, et al. The potential benefit of intracarpal pressure measurement in endoscopic carpal tunnel syndrome surgery--an analysis of EMG findings and pressure values. Acta Medica (Hradec Kralove). 2009;52. doi:10.14712/18059694.2016.106

102. Uchiyama S, Yasutomi T, Momose T, Nakagawa H, Kamimura M, Kato H. Carpal tunnel pressure measurement during two-portal endoscopic carpal tunnel release. Clin Biomech. 2010;25. doi:10.1016/j.clinbiomech.2010.06.019

